# Evidence of a causal and modifiable relationship between kidney function and circulating trimethylamine *N*-oxide with implications for heart and kidney disorders

**DOI:** 10.1101/2022.11.08.22282073

**Authors:** Petros Andrikopoulos, Judith Aron-Wisnewsky, Rima Chakaroun, Antonis Myridakis, Sofia K. Forslund, Trine Nielsen, Solia Adriouch, Bridget Holmes, Julien Chilloux, Sara Vieira-Silva, Gwen Falony, Joe-Elie Salem, Fabrizio Andreelli, Eugeni Belda, Julius Kieswich, Kanta Chechi, Francesc Puig-Castellvi, Mickael Chevalier, Emmanuelle Le Chatelier, Michael T. Olanipekun, Lesley Hoyles, Renato Alves, Gerard Helft, Richard Isnard, Lars Køber, Luis Pedro Coelho, Christine Rouault, Dominique Gauguier, Jens Peter Gøtze, Edi Prifti, MetaCardis Consortium, Jean-Daniel Zucker, Fredrik Bäckhed, Henrik Vestergaard, Torben Hansen, Jean-Michel Oppert, Matthias Blüher, Jens Nielsen, Jeroen Raes, Peer Bork, Muhammad M. Yaqoob, Michael Stumvoll, Oluf Pedersen, S. Dusko Ehrlich, Karine Clément, Marc-Emmanuel Dumas

## Abstract

**Objectives:** The host-microbiota co-metabolite trimethylamine *N*-oxide (TMAO) is linked to increased thrombotic and cardiovascular risks. Here we, sought to i) characterize which host variables contribute to fasting serum TMAO levels in real-life settings ii) identify potential actionable therapeutic means related to circulating TMAO.

**Design:** We applied “explainable” machine learning, univariate-, multivariate- and mediation analyses of fasting plasma TMAO concentration and a multitude of bioclinical phenotypes in 1,741 adult Europeans of the MetaCardis study. We expanded and validated our epidemiological findings in mechanistic studies in human renal fibroblasts and a murine model of kidney fibrosis following TMAO exposure.

**Results:** Next to age, kidney function was the primary variable predicting circulating TMAO in MetaCardis, with microbiota composition and diet playing minor, albeit significant roles. Mediation analysis revealed a causal relationship between TMAO and kidney function decline that strengthened at more severe stages of cardiometabolic disease. We corroborated our findings in preclinical models where TMAO exposure augmented conversion of human renal fibroblasts into myofibroblasts and increased kidney scarring *in vivo*. Mechanistically, TMAO aggravated kidney fibrosis due to ERK1/2 hyperactivation synergistically with TGF-β1 signaling. Consistent with our findings, patients receiving next-generation glucose-lowering drugs with reno-protective properties, had significantly lower circulating TMAO when compared to propensity-score matched control individuals.

**Conclusion:** After age, kidney function is the major determinant of fasting circulating TMAO in adults. Our findings of lower TMAO levels in individuals medicated with reno-protective anti-diabetic drugs suggests a clinically actionable intervention for decreasing TMAO-associated excess cardiovascular risk that merits urgent investigation in human trials.

**Data availability statement:** Raw shotgun sequencing data that support the findings of this study have been deposited in the European Nucleotide Archive with accession codes PRJEB37249, PRJEB38742, PRJEB41311 and PRJEB46098. Serum NMR and urine NMR metabolome data have been uploaded to Metabolights with accession number MTBLS3429; serum GC-MS and isotopically quantified serum metabolites (UPLC–MS/MS) are available from MassIVE with accession numbers MSV000088042 and MSV000088043, respectively.

## Introduction

Over the past two decades, the central role of the commensal gut microbiota in pathologies such as atherosclerosis and type-2 diabetes (T2D) has gained prominence^1^. The microbiota can influence host pathophysiology by producing molecules that directly alter metabolism and/or modulate cellular signaling either locally in the gut or systemically via the circulation^1,2^.

Trimethylamine *N*-oxide (TMAO) is the phase-one liver *N*-oxide of trimethylamine (TMA). TMA is a product of the microbial^3,4,5^ (predominately Firmicutes) metabolism of phosphatidylcholine^6,7^, choline^7^ and *L*-carnitine^8,9,10^, components of the high-fat, high-red meat western diet. TMA is taken up from the gut via the hepatic portal vein and *N*-oxidised into TMAO by host flavin mono-oxygenase 3. High circulating levels of TMAO have been linked to increased thrombotic and cardiovascular risk in animal and human studies, even after adjustment for known cardiovascular risk factors^6,7,8,11^. TMAO is therefore proposed to mediate the higher cardiovascular risk associated with high red meat and fat intake^12^. Fish, the consumption of which is associated with reduced incidence of cardiovascular disease^12^, is also a rich source of TMAO^13^. Dietary TMAO is subject to retro-conversion: *i*.*e*., it can undergo microbial reduction to TMA by *Enterobacteriaceae* followed by hepatic conversion back to TMAO^14^. Concurrent with diet and microbiota composition, TMAO plasma levels also reflect age^15^, sex^16^, kidney function^17,18,19^ and chronic diseases^5,20^. To date, the relative contribution of each of these factors to circulating TMAO levels and, therefore, elevated cardiovascular risk remains unclear. Understanding how serum TMAO levels are regulated in real-life settings could uncover host-TMAO mechanistic targets and identify modifiable and actionable therapeutic factors to lower circulating TMAO levels.

Here, by using a data-driven “explainable” machine learning (ML) strategy^21^, multivariate and univariate analyses of epidemiological data and mechanistic studies in cultured cells and rodents, we sought to objectively identify variables influencing serum TMAO levels in participants of the European multi-center MetaCardis study. Moreover, by taking advantage of the unique cross-sectional MetaCardis design, we queried how variables influencing circulating TMAO manifest at different stages of cardiometabolic disease. Capitalizing on the ML analysis, we aimed to identify (i) novel host TMAO-related mechanistic targets and (ii) a rationale for future interventions that could reduce circulating TMAO levels and thereby decrease associated excess cardiovascular risk **(Suppl.Fig.1)**.

In epidemiological studies, we observed that kidney function is the main modifiable factor consistently regulating fasting serum TMAO levels and our preclinical studies align with the suggestion that elevated circulating TMAO adversely affects kidney function, by increasing kidney fibrotic injury. Interestingly, patients with T2D in the cohort prescribed new-generation antidiabetics (GLP-1 Receptor Agonists^22^; GLP-1RAs) with evidenced reno-protective effects^23^ had lower serum circulating TMAO levels when compared to propensity-score matched controls.

## Results

In a first analysis, we investigated how TMAO levels change depending on disease classification in the MetaCardis population. We confirmed, circulating TMAO significantly increased with cardiometabolic disease severity, in line with previous reports^4,20^ **(Suppl.Fig.2A)**. To identify determinants of circulating TMAO manifesting at prodromal stages of cardiometabolic disease, we focused on the subset of the MetaCardis cohort termed MetaCardis Body Mass Index Spectrum subset (BMIS^23^; N=837) comprising obese/overweight individuals presenting with a range of metabolic syndrome features but not overt T2D or ischaemic heart disease (IHD) **(Suppl.Table.1)**.

### Fasting serum concentration of TMAO is associated with worse cardiometabolic profiles in BMIS MetaCardis population

We explored how TMAO correlated with bioclinical variables related to cardiometabolic health in BMIS individuals. In this group, circulating TMAO was associated with reduced values of estimated glomerular filtration rate (eGFR, Spearman rho=-0.124, pFDR=0.06) and higher fasting concentrations of uric acid (rho=0.114, pFDR=0.09) after adjustment for age, sex, country of recruitment (demographics thereafter) and body mass index (BMI) **(Suppl.Fig.2B, Suppl.Table.2)**. Higher TMAO also positively associated with indicators of central adiposity, including BMI (rho=0.107, pFDR=0.09), visceral body fat rating (rho=0.122, pFDR=0.09) and waist circumference (rho=0.119, pFDR=0.09), after demographics adjustment **(Suppl.Fig.2C, Suppl.Table.3)**. Moreover, BMIS individuals with hypertension (systolic blood pressure over 140mmHg, or receiving therapy for high blood pressure; Methods) had higher plasma TMAO (*P*=0.01, Mann-Whitney test; **Suppl.Fig.2D**). In line with previous studies^13,14,20^, objectively dividing BMIS participants into TMAO clusters using kmeans^25^ revealed that those in the cluster with the highest TMAO levels had consistently worse cardiometabolic traits and were significantly older when compared to those in the cluster with lowest TMAO levels **(Suppl.Fig.2E, Suppl.Table.4)**. Traits of cardiometabolic risk included altered eGFR, elevated liver enzymes and systolic blood pressure **(Suppl.Fig.2F-I)**.

### Age and altered kidney function variables are the main drivers of circulating TMAO in BMIS

To better understand which variables **(Suppl.Table.5)** affect circulating TMAO most, we trained extreme gradient-boosted decision-tree models. We used five-fold cross-validation to predict the Explained Variance (EV) of each variable group on plasma TMAO in the left-out BMIS participants after 100 iterations **(Figure 1A, Suppl.Table.6)**. Microbiota composition alone performed poorly (EV 2%) whilst diet, another purported major contributor to TMAO production, explained less than 5% of TMAO variance. Serum metabolomics, excluding TMAO and its precursor TMA, was the best predictor (EV 12%) with demographics second, explaining 10% of circulating TMAO variance **(Suppl.Table.7)**. The full model, containing all variable groups, accounted, on average, for 18.4% of TMAO variability. The averaged predicted TMAO values by the full model significantly correlated with measured TMAO values (rho=0.473, *P*<2.2×10^−16^; **Suppl.Fig.2J, Suppl.Table.8**).

**Figure 1.**
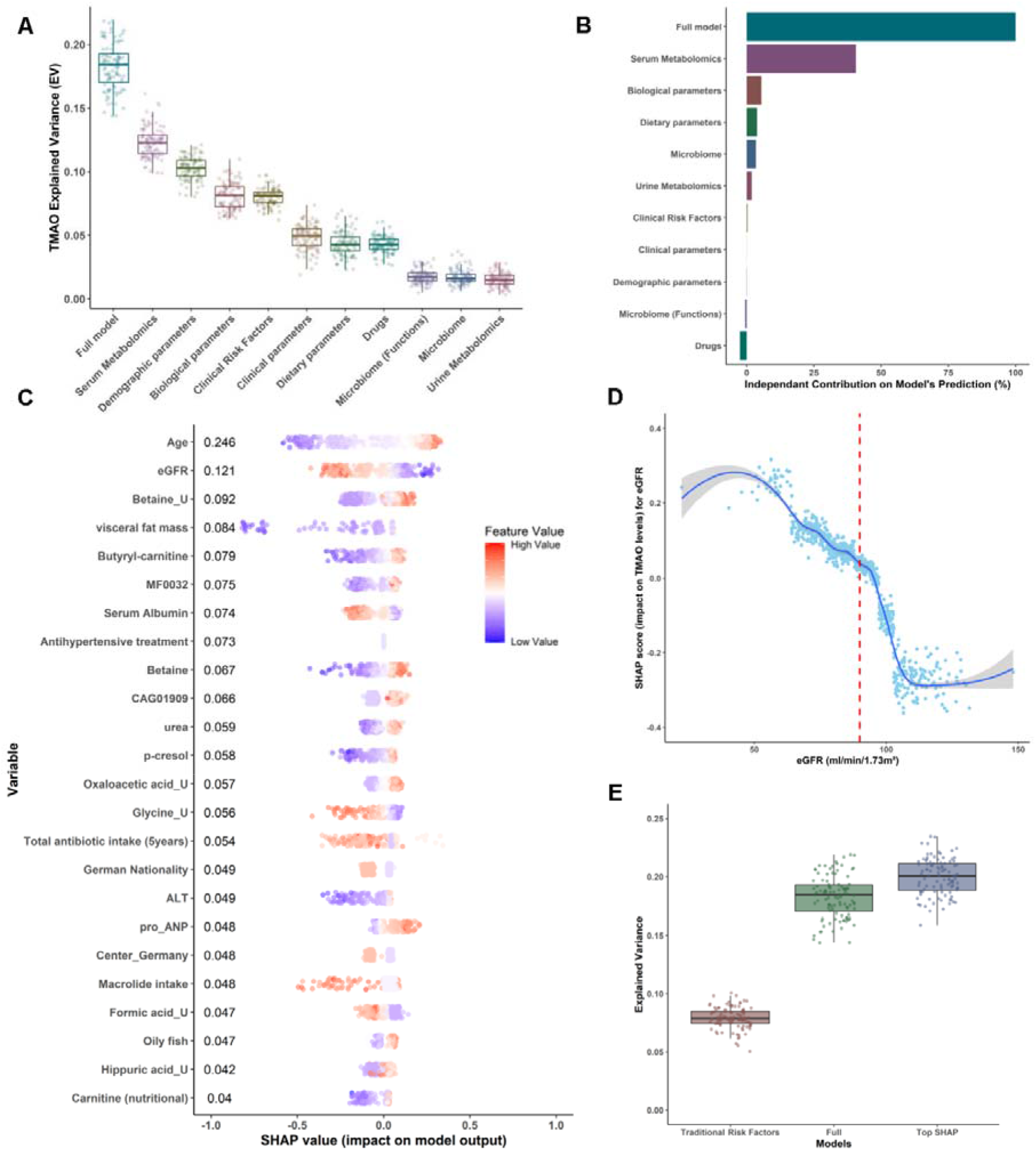
Age and parameters associated with kidney function are the main drivers of circulating TMAO in BMIS MetaCardis participants. **(A)** Coefficients of determination (Explained Variance; EV) of predicted circulating TMAO levels determined by xgboost algorithms after 5-fold cross-validation in the left-out group (**Suppl.Table.6** for N numbers and optimized xgboost parameters per group), trained exclusively on variables from each feature category (**Suppl.Table.5** for a list of variables included in each group), or the full model (containing all variables), after 100 iterations **(Suppl.Table.7). (B)** Averaged independent predictive contribution of each feature category to full model predictions of plasma TMAO, trained as in (A), calculated as the average reduction of the coefficient of determination achieved in relation to the full model (set to 100%) after removing all variables belonging to each feature group after 100 iterations **(Suppl.Table.8**). **(C)** Swarm plots of impact on model output (SHAP values; Suppl.Table.10) for each BMIS individual with complete phenotypic data (N=582) for all variables contributing more than 4% to model predictions of regularized TMAO standard deviation, as determined by xgboost algorithms trained on each feature category. Mean absolute SHAP values from all BMIS participants (N=582) are shown (in descending order) next to each variable. Individual dots, representing each participant, are colored by the inverse-normalized value of the corresponding variable. U denotes urinary metabolites. **(D)** Dependance plot of eGFR values (*x-axis*) *versus* their impact on model outcome (*y-axis*, SHAP values) calculated for each individual in the BMIS cohort (N=837) from algorithms trained exclusively on bioclinical variables **(Suppl.Table.5)**, vertical red line indicates 90mL/min/1.73m^2^. The curve was drawn using locally weighted scatterplot smoothing (LOWESS). **(E)** Boxplots depicting Explained Variance (EV; *R*^2^) of circulating TMAO for BMIS participants computed from algorithms trained on clinical risk factors^29^, the full model containing all variables or all 24 variables contributing more than 4% of regularized TMAO standard deviation to model predictions, as determined by SHAP analysis, after 100 iterations.

We next assessed the independent contribution of each variable group to the predictive power of the full model by training algorithms as above but removing one feature group at a time for 100 iterations. Almost 40% of the explainable variance of the full model (set to 100%) was contributed by serum metabolomic variables, with biological, dietary and microbiota taxonomic variables adding 5.5%, 4% and 3.4%, respectively, independently explained variance **(Figure 1B)**. Other variable categories displayed negligible contribution to prediction suggesting considerable information overlap with the metabolomic, metagenomic, biological and dietary datasets **(Suppl.Table.9)**.

Using feature attribution analysis (SHapley Additive exPlanations; SHAP^21,26^), we assessed how individual variables drive TMAO-predicting models. SHAP analysis identified 24 variables that contributed, more than 4% of the regularized TMAO standard deviation (SD) to model outcomes. Of those, age affected predictions the most, followed by eGFR, urinary betaine, percentage of visceral body fat and serum butyryl-carnitine **(Figure 1C, Suppl.Table.10)**. Besides eGFR, additional variables associated with kidney function including plasma urea^27^, the uremic toxin *p*-cresol^28^ or kidney function decline consequences: i.e. serum albumin^27^ were among those affecting model outcomes the most. This analysis suggests that kidney function is a major determinant of circulating TMAO. The impact of eGFR on model outcomes for BMIS individuals was bimodal with values over 90 mL/min/1.73m^2^, the clinical cut-off value for normal kidney function in adults^25^, predicting reduced plasma TMAO and lower values resulting in increased predicted circulating TMAO **(Figure 1D)**.

We next trained algorithms predicting TMAO using the 24 variables identified by our SHAP analysis (“top SHAP” model) and compared it to models trained by traditional clinical risk factors^29^ or the full model **(Figure 1E)**. The “top SHAP” model significantly (P<2.2×10^−16^, Mann-Whitney test) improved predictions when compared to the full model (EV 21% vs. 18%, on average), presumably by removing noise, supporting the importance of the variables identified by the SHAP analysis.

### Diet and microbiota composition have a modest impact on fasting serum TMAO levels in the BMIS cohort

Our ML models suggested that diet plays a modest role in determining circulating TMAO. To corroborate this, we assessed how consumption of food items rich in TMAO dietary precursors^3^ affected circulating TMAO in BMIS participants with a Food Frequency Questionnaire (FFQ; N=763). After correcting for demographics and BMI, with the exception of oily fish (rho=0.125, pFDR=0.03), circulating TMAO neither significantly associated with habitual consumption of red meat, eggs or milk **(Suppl.Table.11)** nor with the estimated dietary intake of the micronutrients choline, carnitine or betaine **(Suppl.Table.12)**, broadly in agreement with a previous study^3^ **(Figure 2A)**. Additionally, no significant correlations were found between consumption of these food items and the serum levels of TMAO precursors choline, betaine or γ-butyrobetaine^30^. Conversely, circulating TMAO positively correlated with plasma choline, betaine and γ-butyrobetaine, implying an association between the serum concentrations of TMAO precursors.

**Figure 2.**
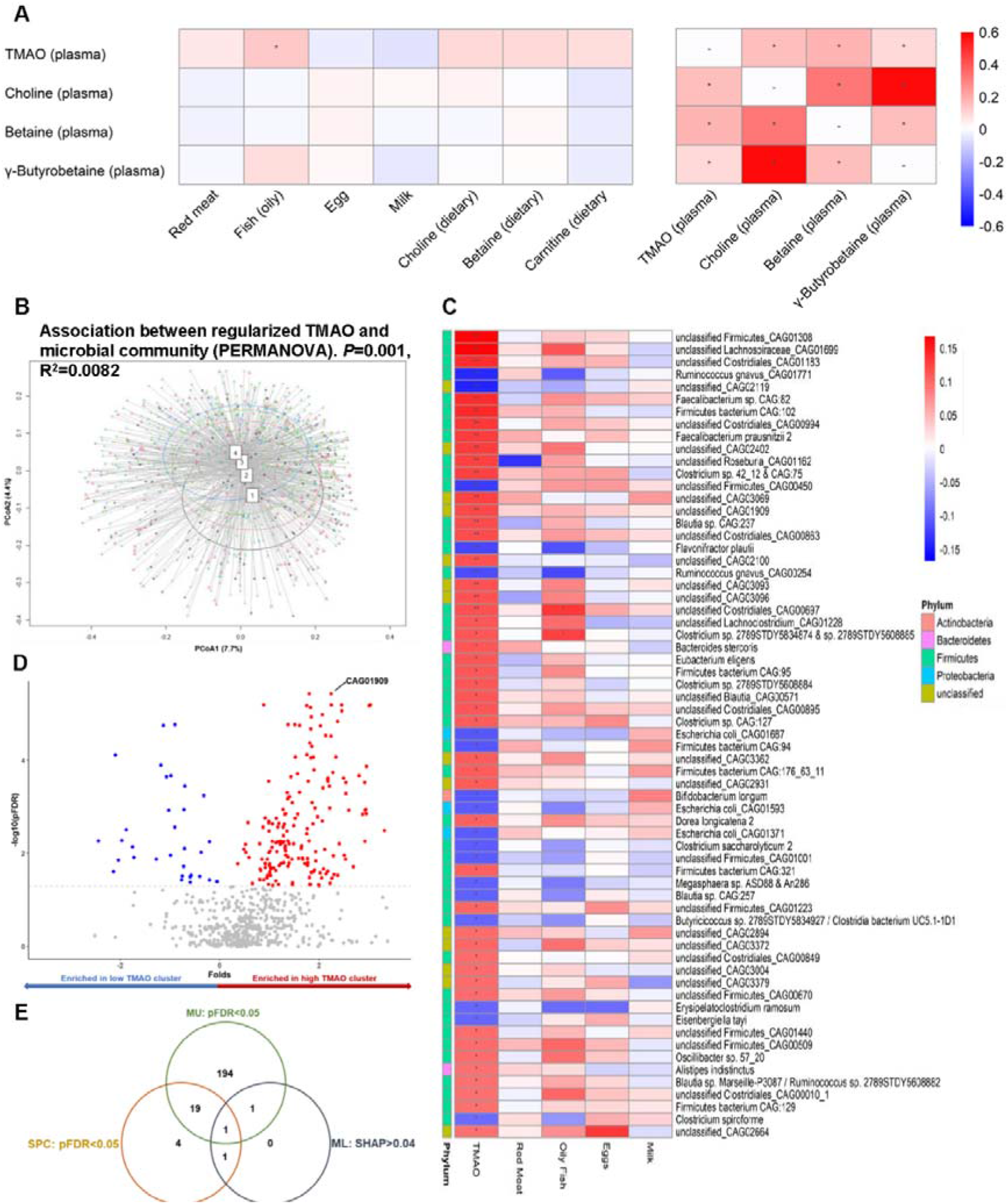
Modest impact of diet and microbiota composition on circulating TMAO in BMIS MetaCardis subjects. **(A)** Associations between circulating TMAO and its precursors with habitual consumption of food items rich in TMAO precursors (N=763; left panel) or with its precursors themselves (right panel) **(Suppl.Table.11, Suppl.Table.12). (B)** Principal Coordinates analysis of Bray-Curtis dissimilarity matrices of participants (N=834) stratified in TMAO clusters by the k-means algorithm (1 the lowest, 4 the highest) at the species level (input; 699 species present in at least 20% of the BMIS population). Insert; PERMANOVA (999 iterations) of taxonomic Bray-Curtis dissimilarity matrices association with regularized TMAO levels with age, sex, and country of recruitment as covariates. **(C)** Overlap of microbiome taxa significantly associated with circulating TMAO (Spearman partial correlations adjusted for age, sex country of recruitment and BMI) and the consumption of food items rich in TMAO precursors in BMIS participants (N=763; **Suppl.Table.13**). **(D)** Volcano plot of differential bacterial species abundances between BMIS participants in the lowest (N=101) and highest (N=147) TMAO clusters (blue; taxa significantly depleted, red; taxa significantly enriched in the high TMAO cluster respectively, pFDR<0.05; **Suppl.Table.14**). (E) Venn diagram summarizing the overlap between taxa associating with circulating TMAO according to our three complimentary analyses (SPC: Spearman correlations; ML: Machine learning and feature attribution analysis; MU: Mann-Whitney U test between high and low TMAO clusters). For all *pFDR<0.05, **pFDR<0. 0.01.

To identify putative microbial taxa influencing serum plasma TMAO, we performed multivariate and univariate analyses. Principal coordinates analysis of Bray-Curtis dissimilarity matrices at the species level for BMIS individuals stratified into TMAO clusters revealed a significant difference in microbiota composition between clusters (*P*=0.033; Figure 2B). Consistent with previous reports^3,31^ and our ML models, multivariate PERMANOVA analysis uncovered a significant, albeit weak, association between circulating TMAO and microbiota composition (*P*=0.001; R^2^ = 0.009; **Figure 2B**) after demographics adjustment. TMAO levels significantly associated with absolute abundance of 65 bacterial species (corrected for bacterial load) in BMIS individuals after correcting for demographics and BMI, primarily (44/65) of the Firmicutes phylum **(Suppl.Table.13, Figure 2C)**. In agreement with Li and colleagues^3^, we did not identify any overlap between bacteria associated with circulating TMAO and higher red meat, milk or eggs consumption in BMIS participants (N=761; **Figure 2C, Suppl.Table.13**). Contrasting low and high TMAO clusters revealed 215 differentially abundant bacterial species between these two groups **(Figure 2D, Suppl.Table.14)**.

We further analyzed the impact on circulating TMAO of the only species, an unknown bacterium taxonomically closely related to ruminoccoci (CAG01909), that contributed at least 4% of the TMAO SD in our prediction models and was significant (pFDR<0.05) for both correlation and differential abundance analyses **(Figure 2E)**. CAG01909 was more abundant and prevalent in the high TMAO cluster when compared to the low **(Suppl.Fig.3A-B)**. Finally, BMIS individuals harboring CAG01909 had significantly higher circulating TMAO than those that lacked it **(Suppl.Fig.3C)**, the CAG1909 abundance significantly correlating with TMAO concentration **(Suppl.Fig.3D)**.

This analysis corroborated the ML models suggesting a small, albeit significant, contribution of microbiota variations on circulating TMAO and identified a bacterium newly associated with higher serum TMAO in BMIS.

### Signatures predicting circulating TMAO shift in different disease groups

To identify putative common variables driving circulating TMAO levels in different disease groups, we trained ML algorithms for T2D (N=561) and IHD sub-cohorts (N=356). Similar to BMIS, microbiota composition and dietary variables alone performed poorly, explaining on average 2% and 1% of TMAO variance, respectively, in T2D and IHD individuals (**Figure 3A and Suppl.Fig.4A** respectively). For the T2D cohort, serum metabolomics was again the best predictor (EV 12.8%) followed by bioclinical (EV 9.6%) and demographic variables (EV 8.7%) with the full model accounting for 16.2% of TMAO variance **(Suppl.Table.17)**. For IHD, individual feature categories, except for serum metabolomics (EV 19.5%) performed poorly, with the full model accounting for 16.1% of circulating TMAO variance **(Suppl.Fig.4A, Suppl.Table.18)**, possibly reflecting reduced power due to smaller sample size and/or IHD group heterogeneity. Feature attribution analysis revealed 19 variables in the T2D cohort and 10 variables in the IHD cohort that contributed more than 4% of regularized TMAO SD to model outcome (**Figure 3B and Suppl.Fig.4B** respectively; **Suppl.Table.19, Suppl.Table.20**). In patients with T2D, model outcome was mostly affected by eGFR followed by age and the serum concentrations of the uremic toxin *p*-cresol^28^ and *D*-threitol, also indicative of kidney function^32^. In patients with IHD, serum butyryl-carnitine, followed by age, alternative healthy eating score (aHEI) related to the ratio of white to red meat intake and the levels of the proinflammatory cytokine IP10, a marker of adverse cardiac remodelling^33^ were the top variables. As in BMIS, the “top SHAP” models significantly improved predictions (EV 24% *versus* 16.2% for T2D; **Figure 3C** and EV 21.3% *versus* 16.1% for IHD; **Suppl.Fig.4C**), when compared to models trained with clinical risk factors or the full model.

**Figure 3.**
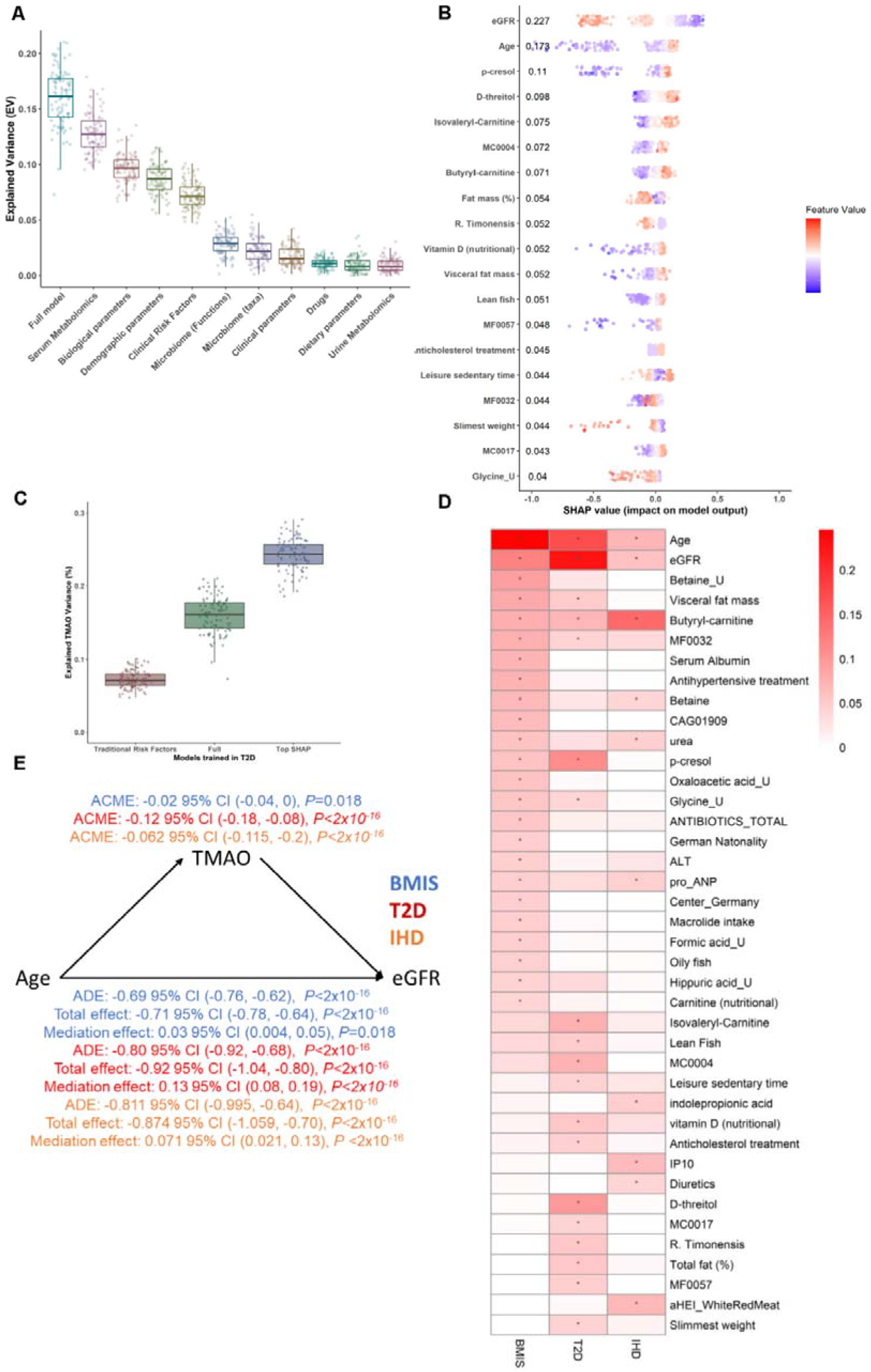
Signatures predicting circulating TMAO shift in different disease groups and TMAO causally mediates eGFR decline with age. **(A)** Explained Variance (EV) of predicted serum TMAO levels **(Suppl.Table.17)** determined by boosted decision trees (Suppl.Table.15 for N numbers and optimized xgboost parameters per variable group), trained exclusively on variables from each variable category (**Suppl.Table.5**, for a list of variables included in each group), or the full model (containing all variables), after 100 iterations in T2D MetaCardis patients. **(B)** Swarm plots of SHAP values (impact on model outcome; **Suppl.Table.19**) for each T2D MetaCardis participant with complete phenotypic data (N=387); represented by individual dots, for all variables contributing more than 4% to model predictions of regularized TMAO standard deviation, computed from xgboost algorithms trained on each feature category. Numbers denote mean absolute SHAP values from all T2D participants (in descending order) next to their corresponding variable. Dots are colored by the inverse-normalized value of their corresponding variable. **(C)** Boxplots depicting Explained Variance (EV; *R*^2^) of circulating TMAO in T2D individuals calculated by algorithms trained on clinical risk factors^31^, the full model containing all variables or all the variables contributing more than 4% of regularized TMAO standard deviation to T2D model predictions, as determined by SHAP analysis, after 100 iterations. **(D)** Heatmap depicting all the variables contributing at least 4% of regularized TMAO standard deviation in model predictions as determined by SHAP analysis in at least one of the MetaCardis disease groups. *Mean absolute SHAP value>0.04. **(E)** Mediation analysis (see Methods) computing the direct effect of TMAO on eGFR decline with age in BMIS (blue), T2D (red) or IHD (orange) MetaCardis participants. ADE: Average direct effect (of Age on eGFR); ACME: average causal mediated effect (of TMAO on eGFR); Total effect: (cumulative effect of age and TMAO on eGFR (ADE + ACME)); Mediation effect: (% of the effect of age on eGFR attributed to TMAO).

Eight variables contributing more than 4% of TMAO SD were shared between BMIS and T2D, including, eGFR, age, and *p*-cresol whilst there were six common features between BMIS and IHD (including eGFR, urea, age and butyryl-carnitine). Only three variables (eGFR, age and butyrylcarnitine) strongly contributed as predictors across all three disease groups **(Figure 3D)**.

We thus identified age and kidney function as the prominent variables influencing fasting circulating TMAO levels. To further substantiate this relationship, we evaluated whether data fit a model where eGFR causally mediates the increase of TMAO concentration with age. Under this model, kidney function achieved significance as mediating ∼20% of the positive association between age and circulating TMAO levels in BMIS with its impact further strengthening in the T2D (55%) and IHD (51%) disease groups **(Suppl.Fig.4D)**, thus corroborating the ML analysis.

### Microbiota composition modestly affects plasma TMAO levels in T2D

In patients with T2D, ML and SHAP analyses revealed an inverse association between fecal *Romboutsia timonensis* abundance and circulating TMAO (Figure 3B), whilst no taxa strongly influenced TMAO predictions in the IHD sub-cohort **(Suppl.Fig.4B)**. *R. timonensis* inversely associated with TMAO concentration (rho=-0.140, *P*=0.0009; **Suppl.Fig.5A**), T2D individuals with detectable *R. timonensis* had significantly lower TMAO levels **(Suppl.Fig.5B)** and *R. timonensis* was depleted in T2D compared to BMIS participants **(Suppl.Fig.5C)**, in line with circulating TMAO **(Suppl.Fig.2A)**. Conversely, in T2D and IHD CAG01909 abundance or presence, unlike BMIS, did not associate with higher circulating TMAO **(Suppl.Fig.5D-5F)**. Further investigation revealed that CAG01909 abundance was inversely associated with metformin intake (rho=-0.141, pFDR=0.042; **Suppl.Fig.5G, Suppl.Table.21**), in accordance with the well-documented effect of drugs on the gut microbiome^34^. Irrespective of TMAO associations with individual species, similarly to BMIS, multivariate PERMANOVA analysis, uncovered a significant, albeit weak, association between circulating TMAO and microbiota composition (P=0.001; R^2^=0.005) after adjustment for demographics.

Collectively, these analyses suggest that the composition of the gut microbiota may only modestly influence circulating levels of TMAO in patients with T2D and in subjects with IHD or T2D abundance of the CAG01909 taxon is not linked with serum TMAO.

### Inference analysis suggests that TMAO may causally mediate the decline of eGFR with age

ML analysis identified age and kidney function as the most prominent variables influencing TMAO. Therefore, we tested the inverse relationship: *i*.*e*., whether TMAO may mediate eGFR decline with age^35^. In BMIS, TMAO modestly, albeit significantly, modulated the inverse relationship between age and eGFR (mediation effect 3%, *P*=0.018), whilst in T2D and IHD the impact of TMAO on kidney function decline with age strengthened (mediation effect 13%, *P*<2.2×10^−16^ and 7.1%, *P*<2.2×10^−16^; **Figure 3E**). Mediation analysis thus indicated that TMAO, far from being a bystander, directly and adversely affects kidney function. Our finding is in agreement with a prospective study reporting that baseline TMAO positively associated with rates of eGFR decline^19^ and with studies in animal models of chronic kidney disease (CKD) where TMAO diet-supplementation increased kidney injury^36,37^ and reduced eGFR.

Interestingly, mediation analysis showed that TMAO’s adverse effect on kidney function strengthened at more severe stages of cardiometabolic disease, implying TMAO synergy with existing pathology (thereby providing “2-hit model”). Accordingly, we next employed preclinical models of kidney injury to gain mechanistic insights into the potential interplay between TMAO and kidney function and the putative molecular nature of the 2^nd^ hit.

### TMAO increases trans-differentiation of human primary renal fibroblasts into myofibroblasts in conjunction with TGF-β1 signaling

Based on TMAO’s detrimental association with kidney damage of diverse aetiologies^17,18^ similarly to renal fibrosis^38^, we investigated the impact of TMAO on human primary renal fibroblasts (HRFs). In HRFs, unlike platelets^39^, TMAO stimulation resulted in rapid [Ca^2+^]_i_ increase **(Figure 4A)**. In endothelial cells the ERK1/2 pathway is activated in a Ca^2+^-dependent manner^40,41^ and ERK1/2 activation exacerbates renal fibrosis^42^. Therefore, we investigated whether a similar pathway operates in HRFs. TMAO challenge increased phospho-ERK1/2 levels in a time-**(Figure 4B)** and dose-dependent **(Figure 4C, Suppl.Fig6A)** manner, at concentrations (10-100μM) relevant to human disease (up to 150μM in patients with CKD)^17^. TMAO-induced ERK1/2 activation was inhibited by a MEK inhibitor, suggesting the activation occurs downstream of Ras-Raf-MEK^42^. Additionally, ERK1/2 activation in response to TMAO was suppressed when intracellular Ca^2+^ was chelated **(Suppl.Fig.6B)** or extracellular Ca^2+^ was removed **(Suppl.Fig.6C)**. Moreover, Ca^2+^ influx was sufficient to activate ERK1/2 in HRFs **(Suppl.Fig.6D);** indicating that TMAO-induced Ca^2+^ influx was required for ERK1/2 activation. We have reported that ERK1/2 phosphorylation was required for HRF trans-differentiation to myofibroblasts in response to TGF-β1^42^. Unlike short-term application (up-to 30min), TMAO stimulation for 48h had minimal effect on ERK1/2 or SMAD3 phosphorylation (**Figure 4D, Suppl.Fig.6E-F)** or on the expression of the myofibroblast marker αSMA **(Figure 4E, Suppl.Fig.6G)**. Conversely, TMAO dose-dependently augmented ERK1/2 pathway activation and myofibroblast trans-differentiation, when co-administered with TGF-β1 in comparison to TGF-β1 or TMAO alone, without affecting SMAD3 phosphorylation **(Figure 4D-E, Suppl.Fig.6E-F)**.

**Figure 4.**
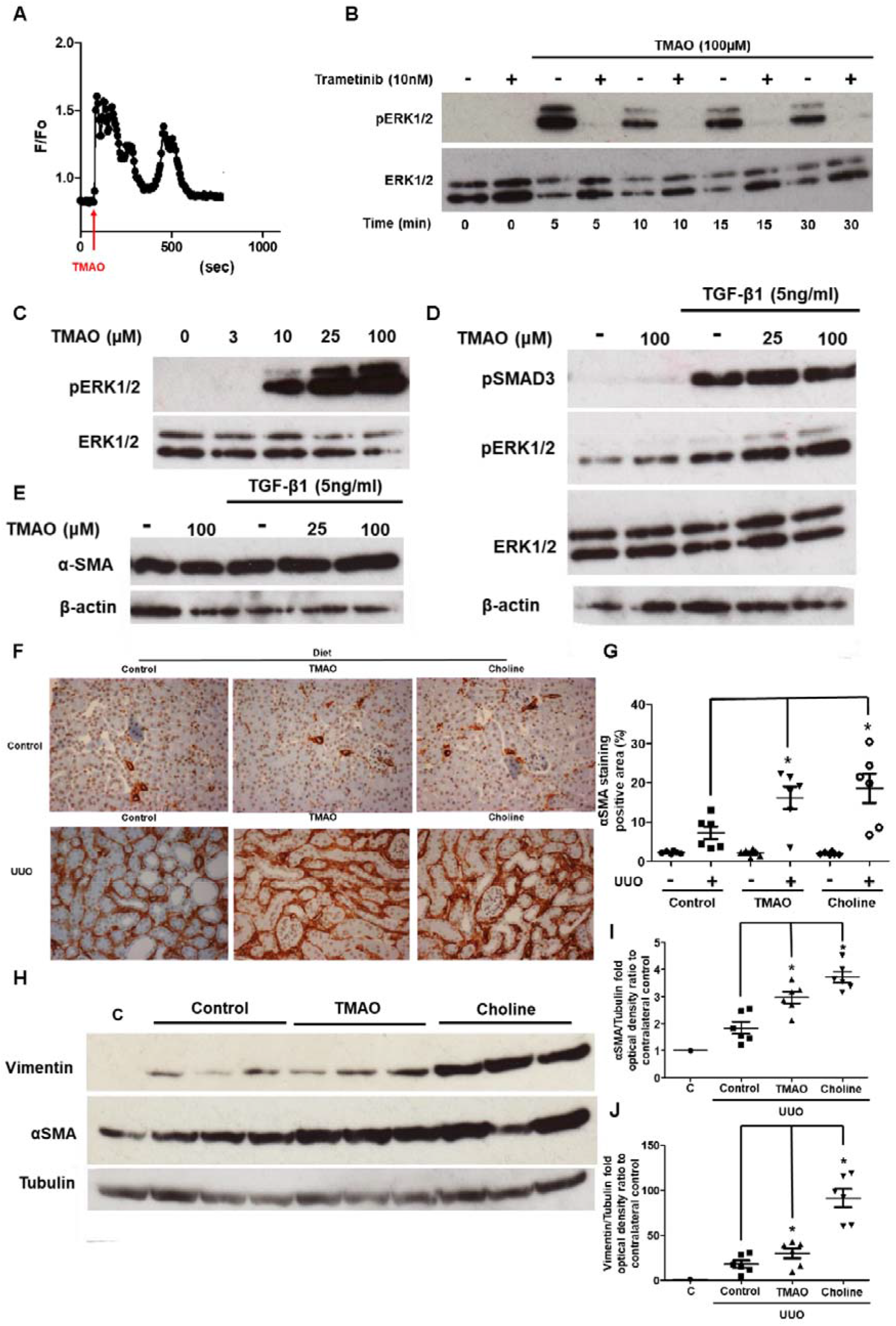
TMAO promotes myofibroblast differentiation and exacerbates renal fibrotic injury. **(A)** Representative ratiometric traces (340/380nm) from Human Renal Fibroblasts (HRFs) loaded with the Ca^2+^ indicator Fura-2 and stimulated with 100μM TMAO. **(B)** Serum-starved HRFs were preincubated with the MEK inhibitor trametinib (10nM) for 30min prior to stimulation with 100μM TMAO for the indicated times. Phospho-ERK1/2 levels were probed by Western blot; membranes were stripped and re-probed for total-ERK1/2. **(C)** Serum-starved HRFs were stimulated with the indicated concentrations of TMAO and phospho-ERK1/2 and total-ERK1/2 levels were determined as in **(B)**. HRFs in complete medium were preincubated with the indicated concentrations of TMAO for 30min and stimulated with TGF-β1 (5nM) or vehicle for 24h. Phospho-ERK1/2, phospho-SMAD3 **(D)** and alpha-smooth muscle actin (αSMA) (E) levels were probed with Western blot. **(F)** Immunostaining with αSMA of kidney sections (20x magnification) from obstructed (UUO; 5days post-surgery) or contralateral sham-operated (control) kidneys. Animals were fed normal chow (control), a diet containing 0.12% w/w TMAO (TMAO) or 1% choline w/w (Choline) for 6weeks prior to surgery, as indicated. N=6 per group. **(G)** Quantification of positive αSMA staining as (%) of stained area/field of view averaged from 5 images per animal. **(H)** Western blot of whole-kidney lysates for αSMA and vimentin expression. Membranes were subsequently stripped and re-probed for tubulin, as loading control. A representative photomicrograph from n=2 Western blots with n=2– 6 animals in each group is shown. OD of the **(I)** αSMA and **(J)** vimentin bands in **(H)** normalized against tubulin. The normalized density of the sham-control samples was arbitrarily set to 1. For all graphs, error bars represent the mean±SEM of data from n=4–6 animals per group. \**P*<0.05 versus the UUO control.

### TMAO increases renal fibrosis: evidence from an intervention in mice

To corroborate our *in vitro* findings suggesting that TMAO directly increases myofibroblast trans-differentiation and establish *in vivo* relevance, we performed Unilateral Ureter Obstruction (UUO) surgery in mice, a model assessing renal fibrosis in the absence of other co-morbidities^43^ that could be also impacted by TMAO. Mice were fed a choline (1% w/w)- or TMAO (0.12% w/w)-supplemented diet for 6 weeks^18^. Subsequently, the ureter of one kidney was ligated whilst the other kidney remained unobstructed **(Suppl.Fig.7A)**. In the unobstructed (control) kidneys αSMA staining was not affected by diet. As expected, five days of UUO resulted in a significant increase in renal αSMA staining of the injured kidney. However, unlike the unobstructed kidneys, TMAO or choline diet supplementation resulted in significantly more myofibroblast expansion (Figure 4F-G). Western blotting of kidney lysates corroborated augmented αSMA and vimentin (another marker of myofibroblasts^42^) expression in injured kidneys of mice receiving TMAO or choline diets **(Figure 4H-J)**. Similar to αSMA, collagen deposition or macrophage infiltration, another indicator of fibrotic kidney damage^38^, were not affected by the TMAO or choline diets in the unobstructed kidneys **(Suppl.Fig.7B-E)**. Conversely, collagen and macrophage staining of kidney slides from UUO kidneys of mice consuming TMAO or choline diets were significantly enhanced compared to controls. Thus, in our experiments in a murine model of kidney fibrosis, TMAO or choline diet supplementation resulted in “hyperactivation” of ERK1/2, mTORC1 and SMAD3 pro-fibrotic signalling^38,42^ **(Suppl.Fig.7F)**.

Collectively, our *in vivo* and *in vitro* findings are consistent with TMAO aggravating kidney fibrosis due to ERK1/2 hyperactivation synergistically probably with activation of the TGF-β1-mediated SMAD3 pathway causing a secondary hit in our disease model.

### Glucagon-Like Peptide-1 Receptor analogues (GLP-1RAs) intake associates with lower serum TMAO concentration in MetaCardis T2D participants

Given the strong bi-directional connection we uncovered between serum TMAO and eGFR, we hypothesized that use of reno-protective medication could be linked with lower circulating TMAO. To identify suitable drugs, we trained algorithms to predict eGFR in MetaCardis patients with T2D, where TMAO appears to have the biggest impact on kidney function, using prescribed medication as input. SHAP analysis revealed that anti-hypertensive and anti-cholesterol treatments had a negative impact on predicted eGFR, probably reflecting more advanced disease **(Figure 5A)**. Conversely, anti-diabetic treatments had a positive effect, with GLP-1RAs being the drugs with the biggest positive impact on predicted eGFR **(Figure 5A, Suppl.Table.22)**. In accordance with their documented reno-protective effect even in T2D patients with no preexisting nephropathy^23^, patients with T2D receiving GLP-1RAs had significantly lower circulating TMAO **(Figure 5B)** than individuals matched for age, sex, hypertension and disease group **(Suppl.Fig.8A-E, Suppl.Table.23)**. This exploratory analysis suggests that GLP-1RAs may reduce serum TMAO concentration and thereby the associated higher risk of IHD.

**Figure 5.**
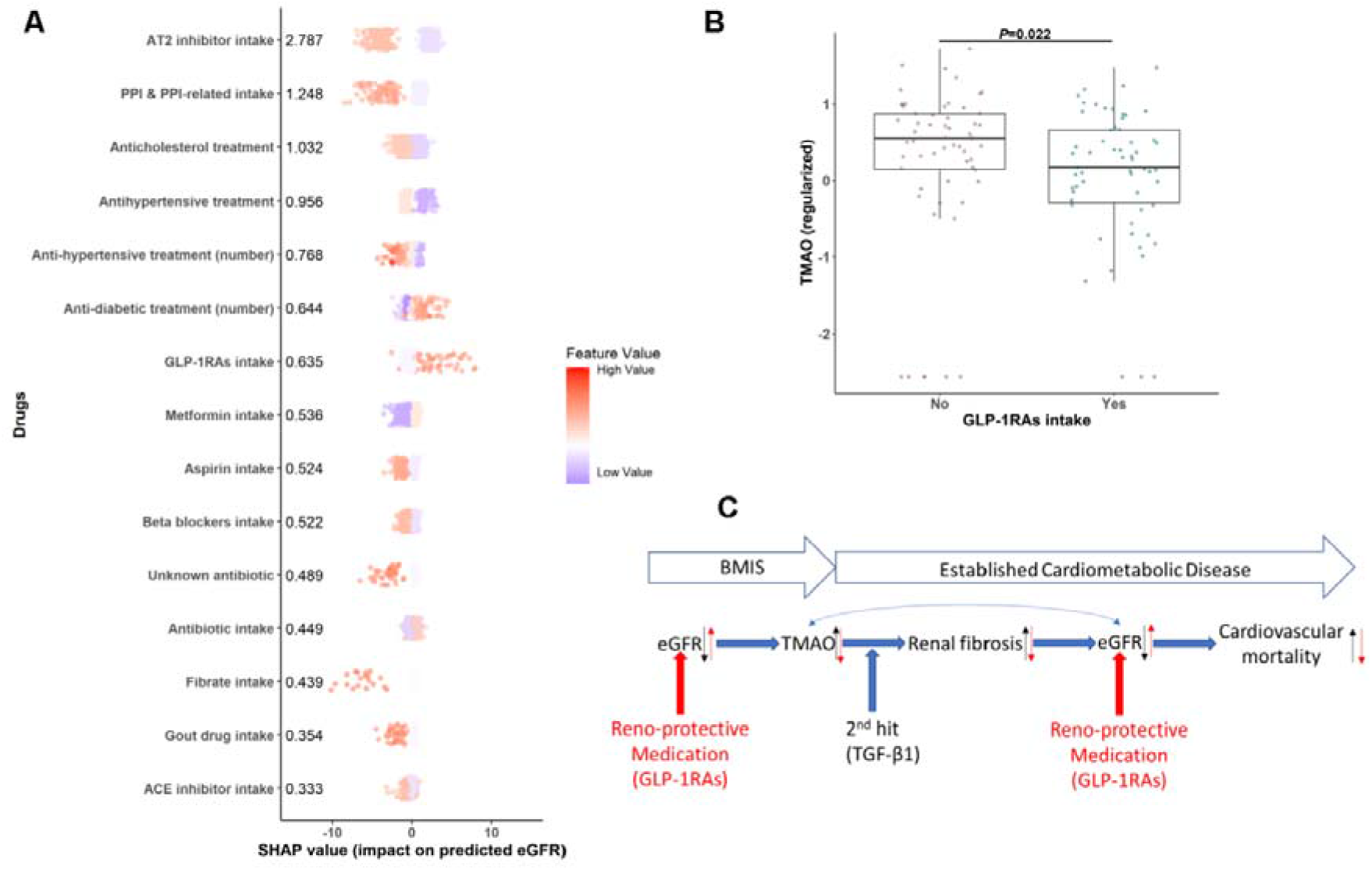
Reno-protective medication is associated with reduced circulating TMAO in MetaCardis participants with T2D. **(A)** Swarm plots of impact on model eGFR predictions (SHAP values; **Suppl.Table.22**) for MetaCardis T2D individuals (N=561) for the top 15 drugs, as determined by xgboost algorithms trained exclusively on prescribed medication. Mean absolute SHAP values from participants with T2D are shown (in descending order) next to each variable. Individual dots, representing each participant, are colored by the inverse-normalized value of the corresponding drug variable. **(B)** Comparison of circulating regularized TMAO levels between subjects with T2D prescribed GLP-1 receptor agonists (GLP-1Ras; N=59) and non-GLP-1Ras treated subjects with T2D propensity-matched for age, sex, BMI, disease group and hypertension status (N=59) (**Suppl.Table.23** for group characteristics). *P*-value determined by Mann-Whitney U test. **(C)** Summary of the main findings of our study. We demonstrate that eGFR, irrespective of disease stage, is the primary modifiable modulator of circulating TMAO. Far from being a bystander, TMAO significantly accelerates the rate of renal output decline by age, with its effect increasing at advanced stages of disease. TMAO promotes renal fibrosis in conjunction with established pathophysiology (two-hit” model) further negatively impacting renal clearance. Accordingly, medication with reno-protective properties (red arrows), such as GLP-1RAs, reduce circulating TMAO levels thereby potentially moderating its adverse effect on kidney function.

## Discussion

With our approach combining epidemiological studies, explorative cellular experiments and murine interventions, we demonstrate that kidney function is the main modifiable factor consistently regulating fasting serum TMAO concentrations and that TMAO adversely impacts on eGFR, at least partially, by increasing kidney scarring synergistically with existing pathophysiology elevating kidney tissue TGF-β1 signaling. Consistent with our findings, use of reno-protective drugs, GLP-1RAs^23^, was associated with lower circulating TMAO in MetaCardis participants with T2D **(Figure 5C)**

Irrespective of cardiometabolic disease severity, gut microbiota composition had a modest (R^2^<0.01), albeit significant, association with serum TMAO levels in the MetaCardis population, in agreement with recent human studies^3,31^. We replicated associations of Firmicutes^3,4,5^ with circulating TMAO and uncovered novel associations between an unknown bacterium (CAG01909) and *R. timonensis*, a marker of less diverse diet in autistic children^44^. However, taxa associated with TMAO diverged between disease modalities. This could at least, partially, be due to medication, suggesting that targeting TMAO production at the species level would be ineffective.

In agreement with a recent study^3^, we did not find any significant association between habitual consumption of red meat and fasting serum levels of TMAO. With a few notable exceptions^3^, the contribution of diet and in particular red meat and *L*-carnitine to TMAO levels, showing an increase following intervention, has predominately been examined in metabolically healthy volunteers^,8,9,10^. In such interventions, often *L*-carnitine has been provided as a dietary supplement, which is poorly absorbed in the small intestine (∼12%), as opposed to dietary *L*-carnitine (∼71%)^45^, and therefore may be more available for microbial catabolism in the upper gut and large intestine, leading to overestimating its role in TMA, and thereby TMAO production. Our observations, similar to the report by Li and colleagues^3^, suggests that in real-life settings where individuals habitually consume meat (75 to 233 g/day in European adults^46^), this contributes minimally to fasting circulating TMAO variability, possibly limiting the isolated effect of dietary manipulation on TMAO levels in real-life, aside strict vegans or vegetarians.

Further, our analyses revealed that the multi-omics signature associated with higher circulating TMAO concentrations and, therefore, elevated cardiovascular risk, shifted as disease progressed with only three variables (age, eGFR and serum butyryl-carnitine) consistently being strong contributors across models built with the BMIS, T2D and IHD disease groups. This is in agreement with our recent report that the majority of markers of dysmetabolism manifest early during cardiometabolic disease development^29^. Accumulation of butyryl-carnitine has been associated with abnormal mitochondrial lipid β-oxidation in T2D^47^, suggesting a link between TMAO and mitochondrial dysfunction consistent with the finding that TMAO binds to protein kinase R-like endoplasmic reticulum kinase and increases mitochondrial stress^48^.

Far from being a bystander, mediation analysis indicated that TMAO significantly accelerated the rate of kidney function decline by age, with its effect increasing in more severe disease stages. Consistent with this, our pre-clinical work uncovered that TMAO primed renal fibroblasts for conversion to myofibroblast, the primary collagen-producing cells in the kidney^38^ and contributed to renal fibrosis, a hallmark of kidney damage irrespective of the underlying cause that significantly contributes to eGFR decline^38^. However, TMAO insult alone was not sufficient to convert renal fibroblasts to myofibroblasts requiring synergy with a pre-existing pathological state, *i*.*e*., pro-fibrotic signaling (“second-hit”) in the form of TGF-β1 stimulation, the most prominent pro-fibrotic cytokine^38^. TGF-β1 expression is increased in ligated UOO mouse kidneys^49^ or kidneys of patients with diabetic nephropathy^50^ and circulating TGF-β1 is elevated in conditions that are risk factors for CKD including hypertension, dyslipidemia and T2D^51^. Therefore, our results suggest that TMAO accelerates eGFR decline in concert with existing pro-fibrotic signaling (i.e. TGF-β1; “second-hit”), consistent with observations in animal models of CKD^36,37^ and humans^18,19^.

Supporting our assertion that TMAO accelerates cardiovascular disease progression together with existing pathology, at least partially by impacting the kidney, TMAO increased all-cause mortality only in individuals with eGFR<90 mL/min/1.73m2^52^ and circulating TMAO levels in healthy adults were not indicative of future atherosclerotic burden^53^.

TMAO is an independent risk factor of cardiovascular morbidity and mortality in patients with established IHD or CKD^6,17,18^ with accumulating evidence suggesting direct causal effects^40^. Our work suggests that reno-protective strategies could potentially lower circulating TMAO and therefore preserve kidney function in individuals with high circulating TMAO in the presence of risk factors known to increase TGF-β1 signaling (i.e hypertension and T2D^51^). Indeed, we observed that patients with T2D prescribed GLP-1RAs, drugs with documented reno-protective and beneficial cardiovascular effects^22,23^, had significantly lower fasting plasma TMAO levels than propensity-score matched controls.

## Conclusion

Collectively, our findings demonstrate that eGFR, irrespective of disease stage, is the primary modifiable modulator of circulating TMAO, which then by promoting renal fibrosis in conjunction with established pathophysiology (“second-hit” model) further negatively impacts kidney function. Accordingly, we observe that intake of GLP-1RAs is associated with lower circulating TMAO levels, thereby potentially moderating the adverse effect of TMAO on kidney function and suggesting a putative mechanism for these drugs observed reno-protection in large pharmaceutical trials^23^.

Our work, conceptually advances understanding of how TMAO levels (and therefore associated cardiovascular risk) are regulated in humans with a wide range of Cardiometabolic disease burden in real-life settings. Additionally, we uncover a direct mechanistic link between TMAO and renal fibrosis in conjunction with existing co-morbidities (known to elevate TGF-β1 signaling) such as hypertension and T2D. Furthermore, our findings suggest that therapeutic modalities preserving kidney function could markedly benefit and reduce cardiovascular risk in individuals with high circulating TMAO in the presence of risk factors (T2D, hypertension or metabolic syndrome). This merits urgent testing in a longitudinal independent clinical trial.

## Data Availability

Raw shotgun sequencing data that support the findings of this study have been deposited in the European Nucleotide Archive with accession codes PRJEB37249, PRJEB38742, PRJEB41311 and PRJEB46098. Serum NMR and urine NMR metabolome data have been uploaded to Metabolights with accession number MTBLS3429; serum GC-MS and isotopically quantified serum metabolites (UPLC-MS/MS) are available from MassIVE with accession numbers MSV000088042 and MSV000088043, respectively.

## Author contribution

P.A., K.C. and M.E.-D. developed the present project concept and protocol. K.C. (coordinator), O.P., M.S., S.D.E., P.B., J.R., M.-E.D., F.B. and J.N. conceived the overall objectives and study design of the MetaCardis initiative. MetaCardis cohort recruitment, phenotyping and lifestyle: J.A.-W., R.C., T.N., J.-E.S., F.A, L.K., H.V., T.H., J.-M.O. and M.B. and supervised by K.C., M.S. and O.P. Data curation: R.C., S.K.F., J.A.-W. and T.N. Bacterial cell count measurement: S.V.S. and G.F. Serum and urine metabolome profiling: A.M., J.C., M.T.O. and L.H. Biochemical analyses: J.-P.G. Bioinformatics and statistical analyses: P.A., S.K.F., G.F, S.V.S., R.A., E.L.C., L.P.C., E.P. and E.B. Dietary FQQ analyses: S.A., and B.H. Modelling of phenotypic data: P.A. Cellular in vitro experiments: P.A. Murine in vivo experiments P.A. and J.K. with supervision from M.M.Y. The manuscript was written by P.A., KC and M-E.D. with input from J.-A.W., R.C., S.K.F., M.M.Y., M.S., O.P. and S.D.E. All authors participated in project development, discussion of results and revision of the article and approved the final version for submission.

## Acknowledgements

We thank the subjects for their participation in the MetaCardis study and particularly patient associations (Alliance du Coeur and CNAO) for their input and interface, as well as Dr Dominique Bonnefont-Rousselot (Department of Metabolic Biochemistry, Pitié-Salpêtrière hospital) for the analysis of plasma lipid profiles. We thank the nurses, technicians, clinical research assistants and data managers from the Clinical investigation platform at the Institute of Cardiometabolism and Nutrition for patient investigations, at the CRNH (Centre de recherche en Nutrition Humaine CRNH-Ile de France) and, the Clinical Investigation Center (CIC) from Pitié-Salpêtrière Hospital for investigation of healthy controls. Quanta Medical provided regulatory oversight of the clinical study and contributed to the processing and management of electronic data. Similarly, we are indebted to the MetaCardis consortium (http://www.metacardis.net/) collaborators for contributions at multiple levels since the consortium start in 2012. A full list of collaborators is given in the Supplementary Section.

## Main Figures

## Supplemental Figures

**Supplemental Figure 1.**
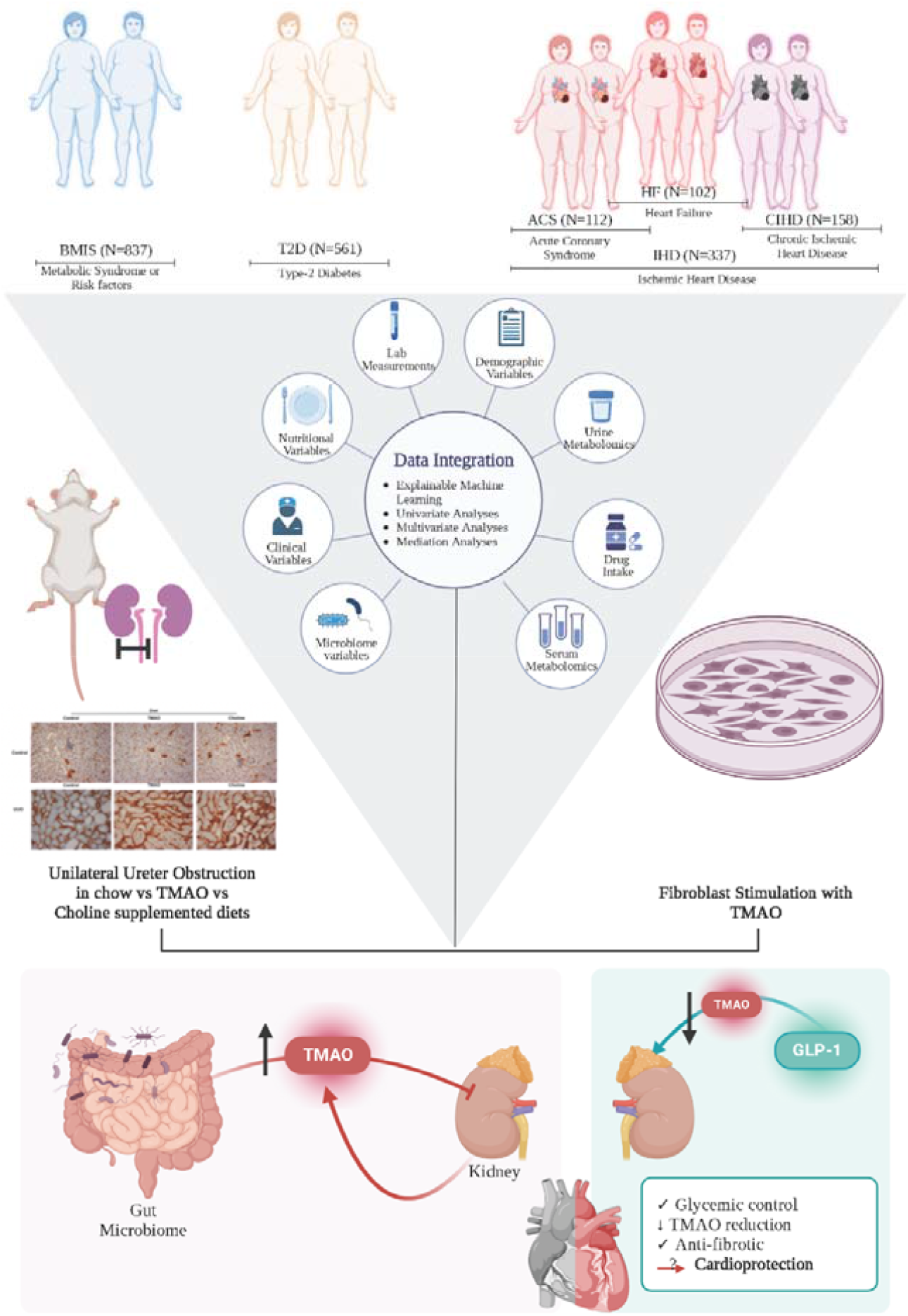
Overview of study design and aims. Here we used an integrated approach comprising Machine Learning (ML), multivariate, univariate and mediation analyses to objectively characterize which host parameters contribute to plasma TMAO levels in the multi-center European MetaCardis study. Capitalizing on the bioinformatics analysis we aimed to i) uncover novel host-TMAO mechanistic targets and ii) identify actionable approaches (*e*.*g*. precision nutrition and/or pharmacology interventions) to reduce systemic TMAO levels.

**Supplemental Figure 2.**
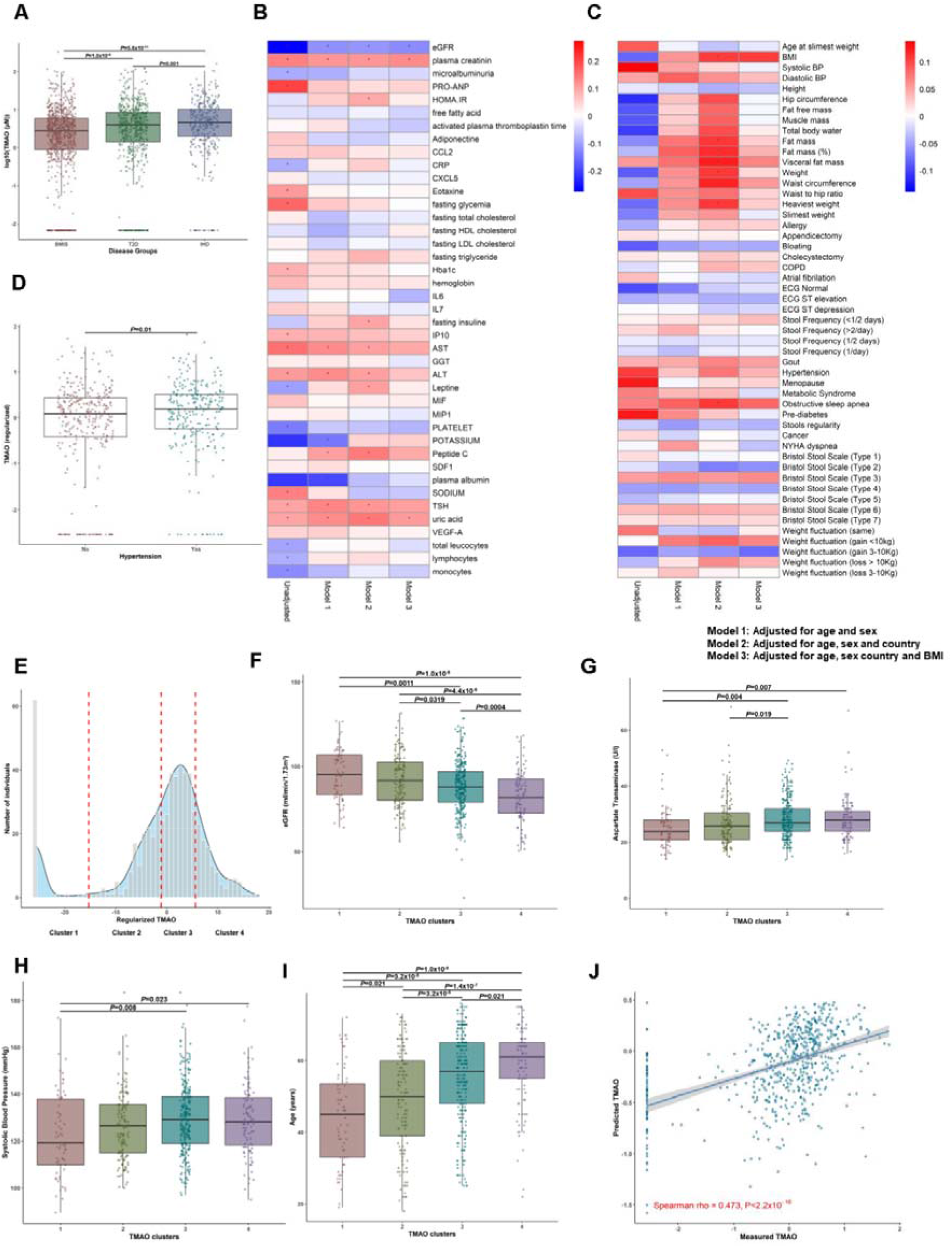
TMAO is associated with worse cardiometabolic profiles in study participants with metabolic syndrome-related risk factors or pathologies but not over cardiometabolic disease (BMIS subgroup N=581). **(A)** Boxplot illustrating differences in fasting circulating TMAO levels in the three MetaCardis patient groups (**Suppl.Table.1** for sub-cohort characteristics). *P* determined by Mann-Whitney U test. **(B)** Spearman correlations between circulating TMAO levels and bioclinical variables unadjusted, adjusted for age and sex (Model 1), age sex and country of recruitment (Model 2) or age, sex country of recruitment and BMI (Model 3), *pFDR<0.1. **(Suppl.Table.2). (C)** Spearman correlations between plasma TMAO and clinical variables, corrected as in **(A), (Suppl.Table.3). (D)** Comparison of circulating TMAO levels between BMIS participants classed as hypertensives, *P* determined by Mann-Whitney U test. **(E)** Density plot illustrating the distribution of regularized circulating TMAO levels in BMIS (N=582). The population was split into four TMAO clusters using the k-means algorithm. Dashed red lines denote the cut-off values for this distribution. Between group comparisons of selected variables for BMIS participants split into clusters according to their circulating TMAO levels for eGFR (**F**; mL/min/1.73m^2^), aspartate transaminase (**G**; U/l), systolic blood pressure (**H**; mmHg) and age at the time of recruitment (**I**, years). *P* values were determined with pairwise Mann-Whitney U tests corrected for multiple comparisons with the Benjamini-Hochberg method. **(J)** Predicted (averaged after 100 iterations; y-axis) regularized plasma TMAO of BMIS participants by the BMIS-trained full-model *versus* actual measured regularized TMAO values (*x-axis*) for BMIS individuals (N=582) **(Suppl.Table.8);** insert Spearman rho and *P*-value of predicted *versus* measured values.

**Supplemental Figure 3.**
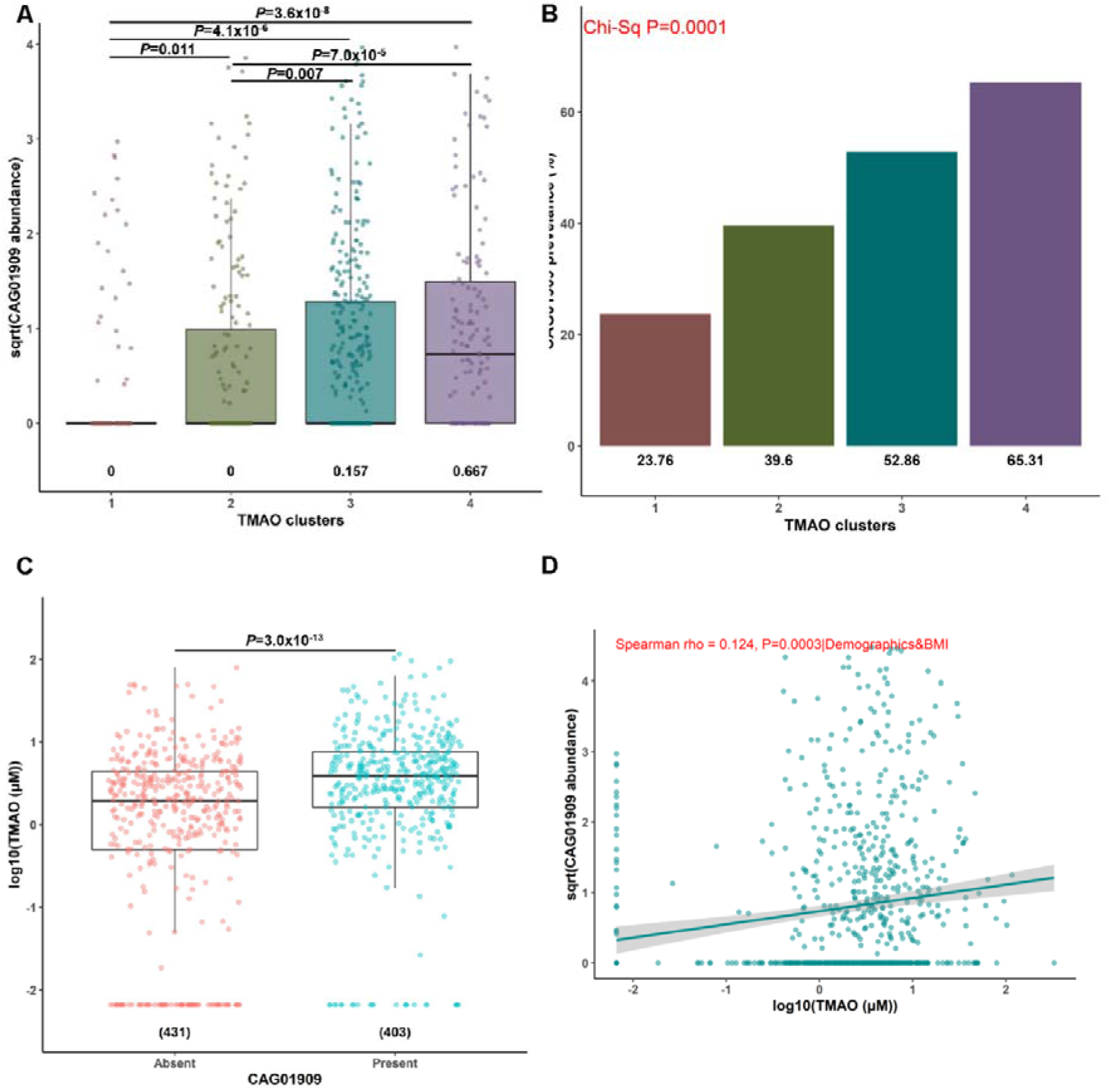
CAG01909, associates with higher circulating TMAO in BMIS (N=834) individuals. Comparisons of normalized to bacterial load expression levels (square-root transformed) of **(A)** CAG01909, an unknown bacterium in BMIS participants divided into clusters according to circulating TMAO levels with the k-means algorithm. Median abundance level for each cluster is shown below the corresponding box plot, all pairwise comparisons performed with Mann-Whitney U tests (corrected for multiple testing with the Benjamini-Hochberg method). **(B)** Prevalence ((%); in parentheses under corresponding boxplots) of *CAG01909* in BMIS participants (N=834) split into TMAO clusters as in (A). *P* value determined by Chi-square test. **(C)** BMIS participants were split into those where CAG01909 is present in their gut microbiota (N=401) and those where it is absent (N=433) and circulating log-transformed TMAO levels between the two groups were compared (Mann-Whitney U test). **(D)** Linear-regression-based scatterplot showing correlation between normalized by bacterial load CAG01909 abundance levels (square-root transformed for visualization purposes) and TMAO (log_10_-transformed). Adjusted (age, sex, country of recruitment and BMI) Spearman rho=0.123 and pFDR=0.032 for CAG01909 **(Suppl.Table.13)**.

**Supplemental Figure 4.**
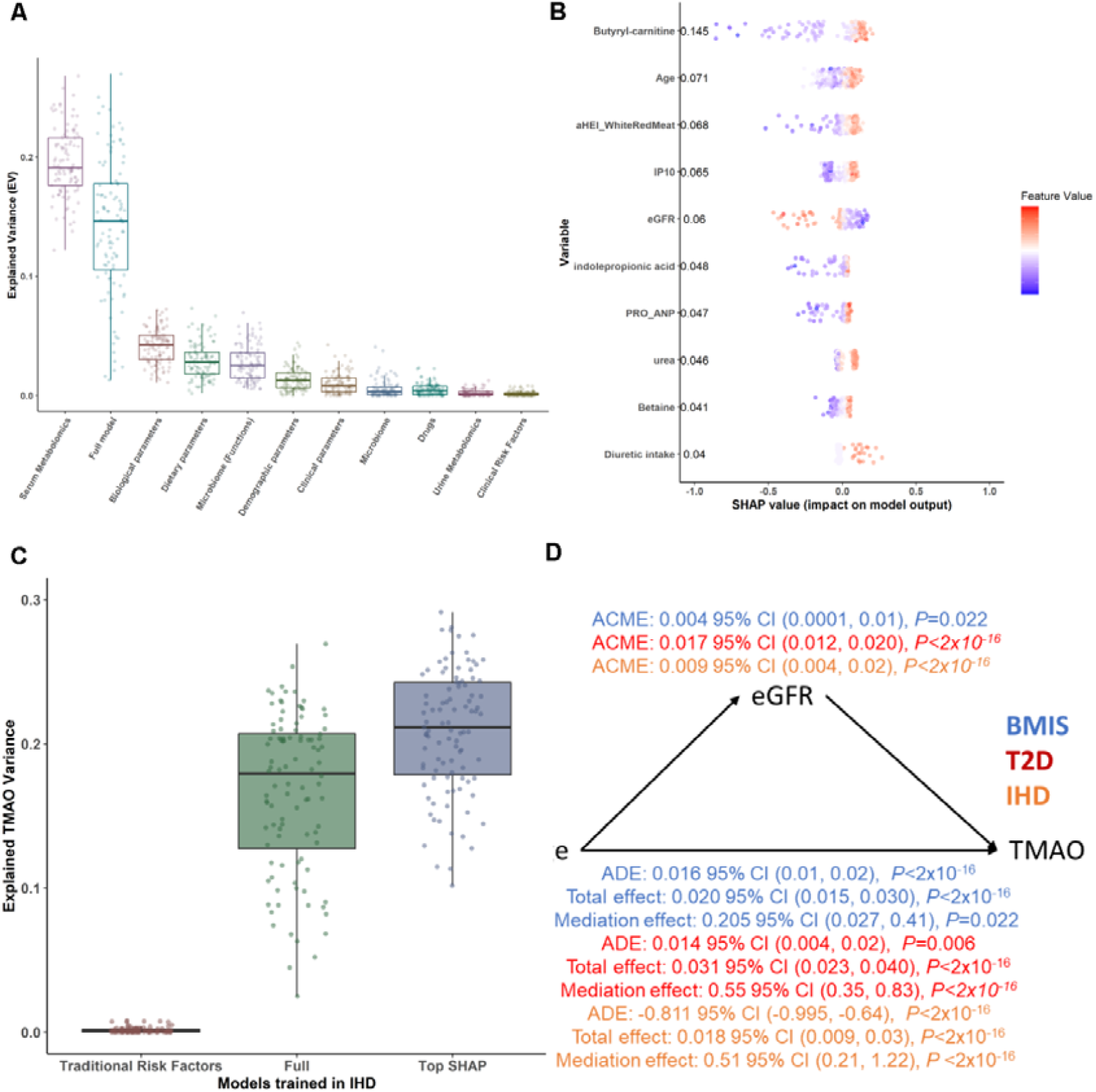
Boosted decision tree algorithms predict circulating TMAO in IHD MetaCardis participants. **(A)** Explained Variance (EV) of predicted serum TMAO levels **(Suppl.Table.18)** determined by boosted decision trees (methods), trained exclusively on variables from each feature category (**Suppl.Table.5** for a list of variables included in each group), or the full model (containing all variables), after 100 iterations (**Suppl.Table.16** for N numbers and optimized xgboost parameters per variable group) in IHD MetaCardis individuals. **(B)** Swarm plots of SHAP values (impact on model outcome; *x-axis*) for each IHD MetaCardis participant with complete phenotypic data (N=221), for all variables contributing to model predictions more than 4% of regularized TMAO standard deviation, computed from xgboost algorithms trained on each feature category. Numbers denote mean absolute SHAP values from all IHD participants (in descending order) next to their corresponding variable **(Suppl.Table.20)**. Dots, representing IHD individuals, are colored by the inverse-normalized value of their corresponding variable. **(C)** Boxplots depicting Explained Variance (EV; *R*^2^) of circulating TMAO in IHD individuals computed by algorithms trained on clinical risk factors^29^, the full model containing all variables or all the variables contributing more than 4% of regularized TMAO standard deviation to IHD model predictions, as determined by SHAP analysis, after 100 iterations. **(D)** Mediation analysis computing the direct effect of eGFR on TMAO increase with age in BMIS (blue), T2D (red) or IHD (orange) MetaCardis participants. ADE: Average direct effect (of Age on TMAO); ACME: average mediation effect (of eGFR on TMAO); Total effect: (cumulative effect of age and eGFR on TMAO (ADE + ACME)); Mediation effect: (% of the effect of age on circulating TMAO attributed to eGFR).

**Supplemental Figure 5.**
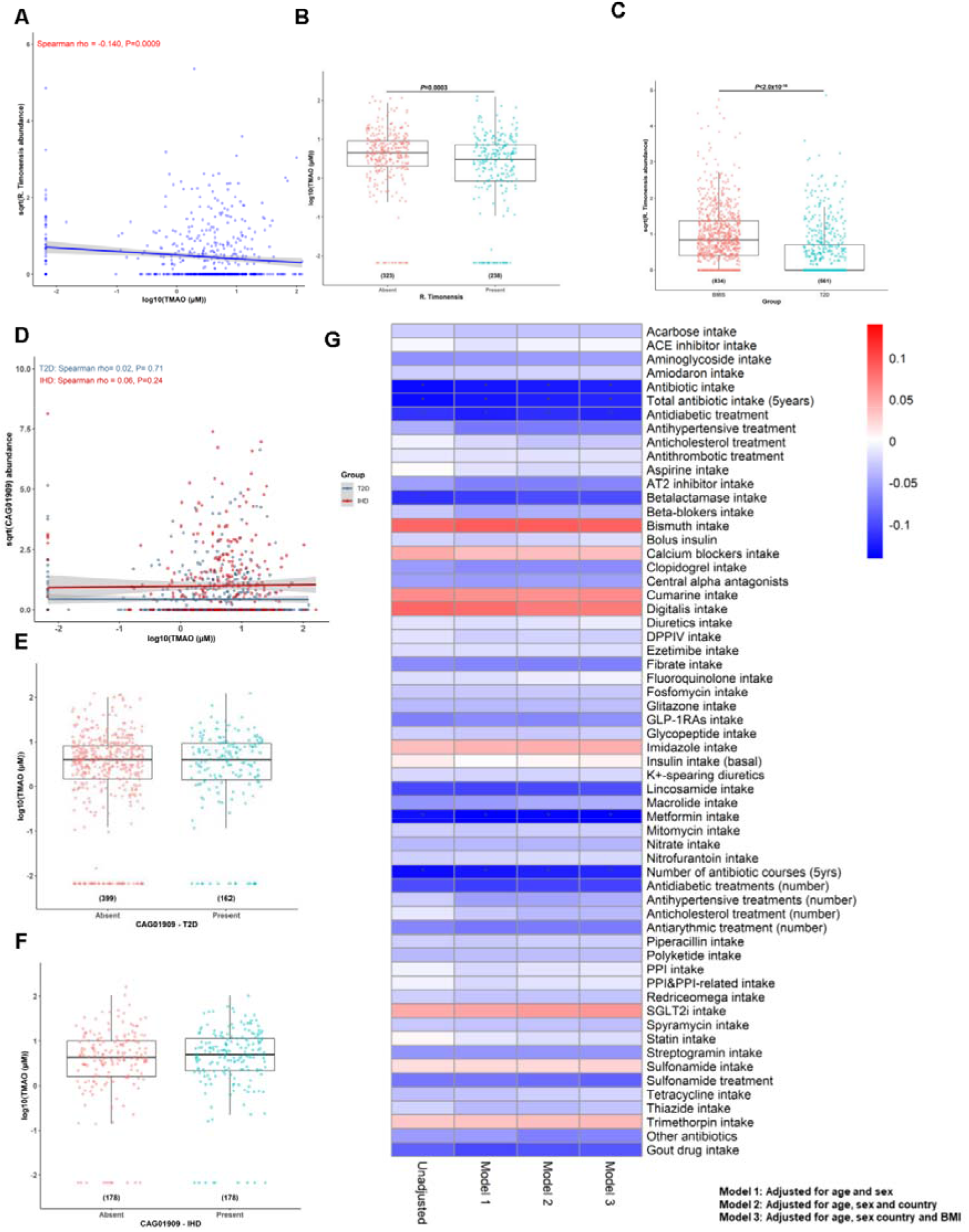
In MetaCardis T2D participants *R. timonensis* inversely associates with circulating TMAO whilst BMIS-identified CAG01909 does not in overt disease. **(A)** Linear-regression-based scatterplot showing associations between normalized by bacterial load *R. timonensis* abundance levels (square-root transformed for visualization purposes) and TMAO (log_10_-transformed). Unadjusted spearman rho=-0.140 and P=0.009. **(B)** Comparison of log-transformed circulating TMAO between T2D individuals with detectable (present) and undetectable (absent) fecal *R. timonensis* (Mann-Whitney test). Number of individuals in each group shown in parenthesis. **(C)** Boxplot illustrating differences in *R. timonensis* fecal abundance in the BMIS and T2D MetaCardis patient groups (group sizes in parentheses), *P* determined by Mann-Whitney U test. **(D)** Linear-regression-based scatterplot showing correlation between normalized by bacterial load CAG01909 expression levels (square-root transformed for visualization purposes) and TMAO (log_10_-transformed) in T2D and IHD MetaCardis subjects. Comparison of log-transformed circulating TMAO between T2D **(E)** and IHD **(F)** MetaCardis subjects with detectable (present) or undetectable (absent) fecal CAG01909 (Mann-Whitney test). Number of individuals in each group shown in parenthesis. **(G)** Heatmap illustrating Spearman correlations between bacterial load-normalized abundance of CAG01909 and intake of medication in T2D MetaCardis individuals (N=561) unadjusted, adjusted for age and sex (Model 1), age sex and country of recruitment (Model 2) or age, sex country of recruitment and BMI (Model 3), *pFDR<0.1. **(Suppl.Table.21)**.

**Supplemental Figure 6.**
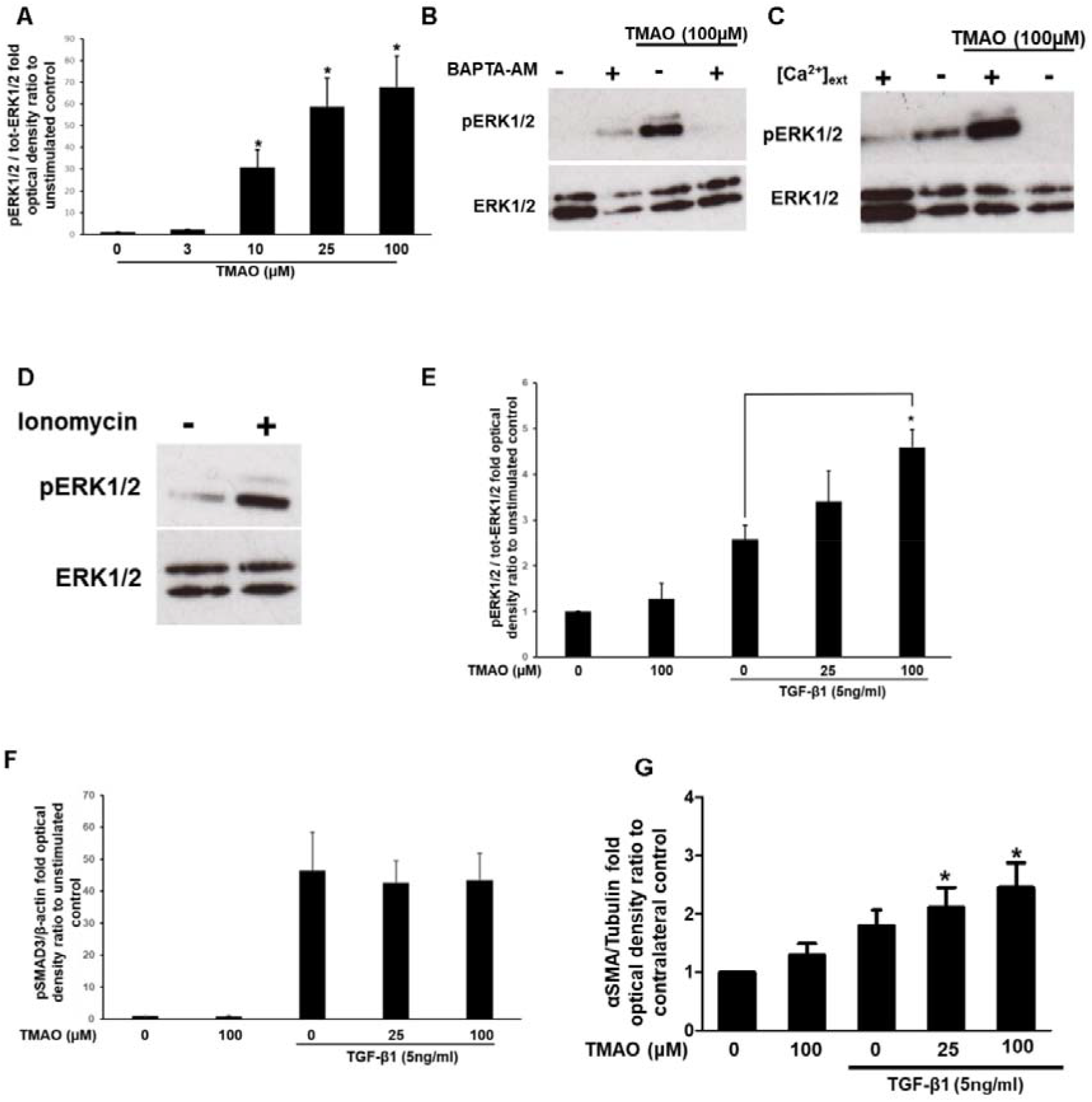
TMAO activates ERK1/2 via a Ca^2+^-sensitive pathway in human renal fibroblasts. **(A)** Optical density (OD) of pERK1/2 levels normalized against total ERK1/2 for experiments represented in **Figure 4C**. The normalized density of the control unstimulated samples was arbitrarily set to 1. **(B)** Serum-starved Human Renal Fibroblasts (HRFs) were loaded with the Ca^2+^ chelator BAPTA-AM (20μM; 20min) and then stimulated with TMAO (100μM) for 10min. ERK1/2 activation was probed by Western blot and membranes were stripped and re-probed for total ERK1/2, a representative image of N=3 independent experiments is shown. **(C)** HRFs were serum-starved in complete physiological medium for 45min, the medium was aspirated and cells were incubated for further 15min in physiological medium with Ca^2+^ omitted from the buffer. Subsequently, cells were stimulated with 100μM TMAO for 10min and ERK1/2 activation was probed as in **(B). (D)** Serum-starved HRFs were stimulated with the Ca^2+^ ionophore ionomycin (100μM) and phospho-ERK1/2 levels were probed as in (B). OD of pERK1/2 (E) or pSMAD3 (F) normalized by total ERK1/2 or β-actin respectively for experiments represented in **Figure 4D**. The normalized density of the control unstimulated samples was arbitrarily set to 1. (G) OD of αSMA normalized by β-actin levels for experiments represented in **Figure 4E**. For all N=3, error bars represent ±SEM.

**Supplemental Figure 7.**
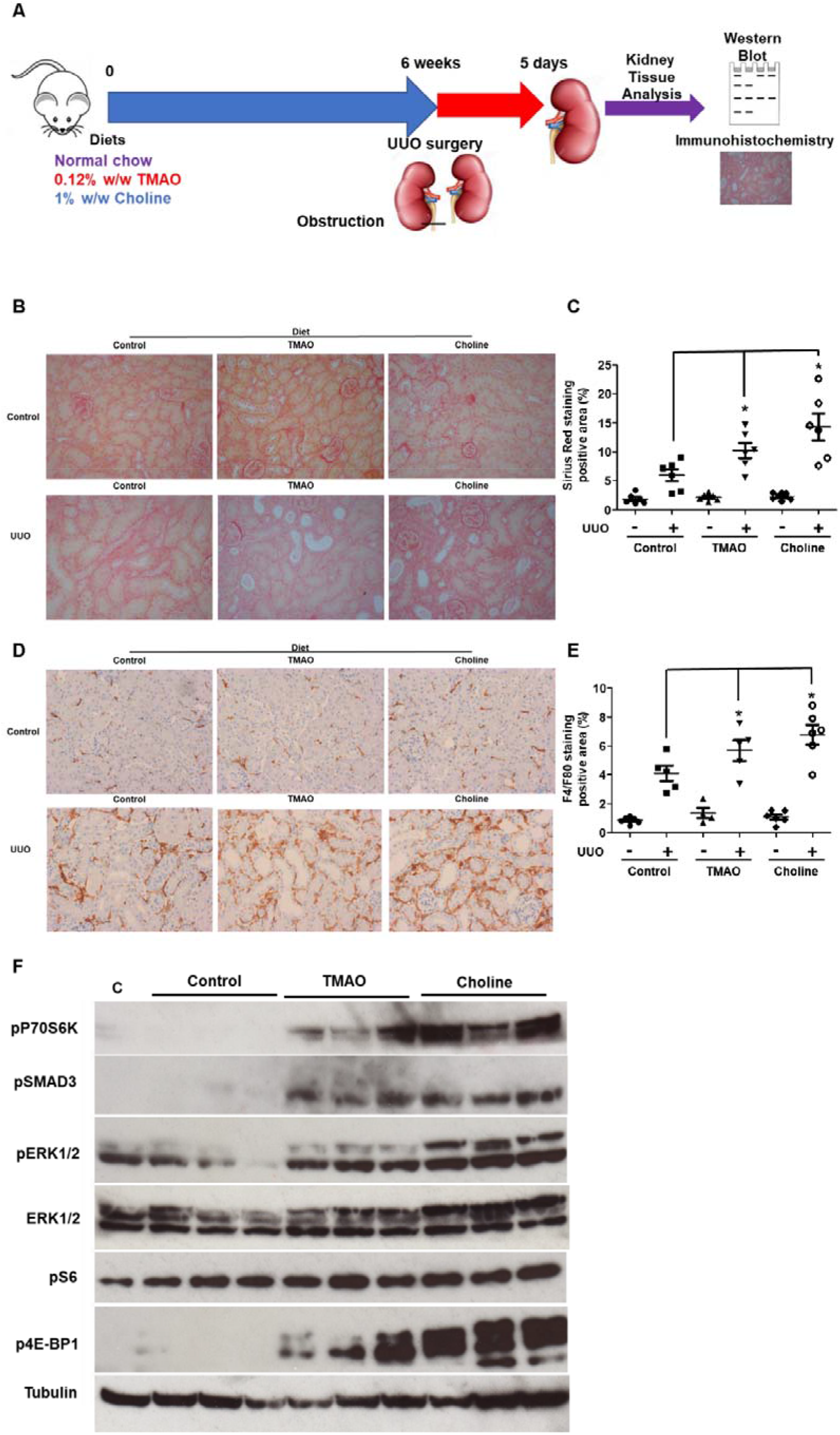
TMAO or choline diet exacerbates collagen deposition, macrophage infiltration and activate pro-fibrotic signaling in kidneys of mice that underwent Unilateral Ureter Obstruction (UUO) surgery. **(A)** Outline of the animal experiments. **(B)** Sirius red staining of kidney sections (20x magnification) from obstructed (UUO; 5days post-surgery) or contralateral sham-operated (control) kidneys. Animals were fed normal chow (control), a diet containing 0.12% w/w TMAO (TMAO) or 1% choline w/w (Choline) for 6weeks prior to surgery, as indicated. N=6 per group. **(C)** Quantification of collagen deposition staining as (%) of positive Sirius red area/field of view averaged from 5 images per animal. **(D)** Immunostaining of kidney sections (20x magnification) from UUO or control kidneys as in **(B)** with the macrophage marker F4/F80. **(E)** Quantification of macrophage infiltration as % of F4/F80 staining/ field of view of images from (D); N=6 animals/group. **(F)** Western blot for profibrotic signaling pathways activation from UUO kidneys from animals treated as in **(A)**. C: contralateral kidney from a control-diet fed animal; TMAO: 0.12% w/w TMAO diet; Choline: 1% w/w Choline diet.

**Supplemental Figure 8.**
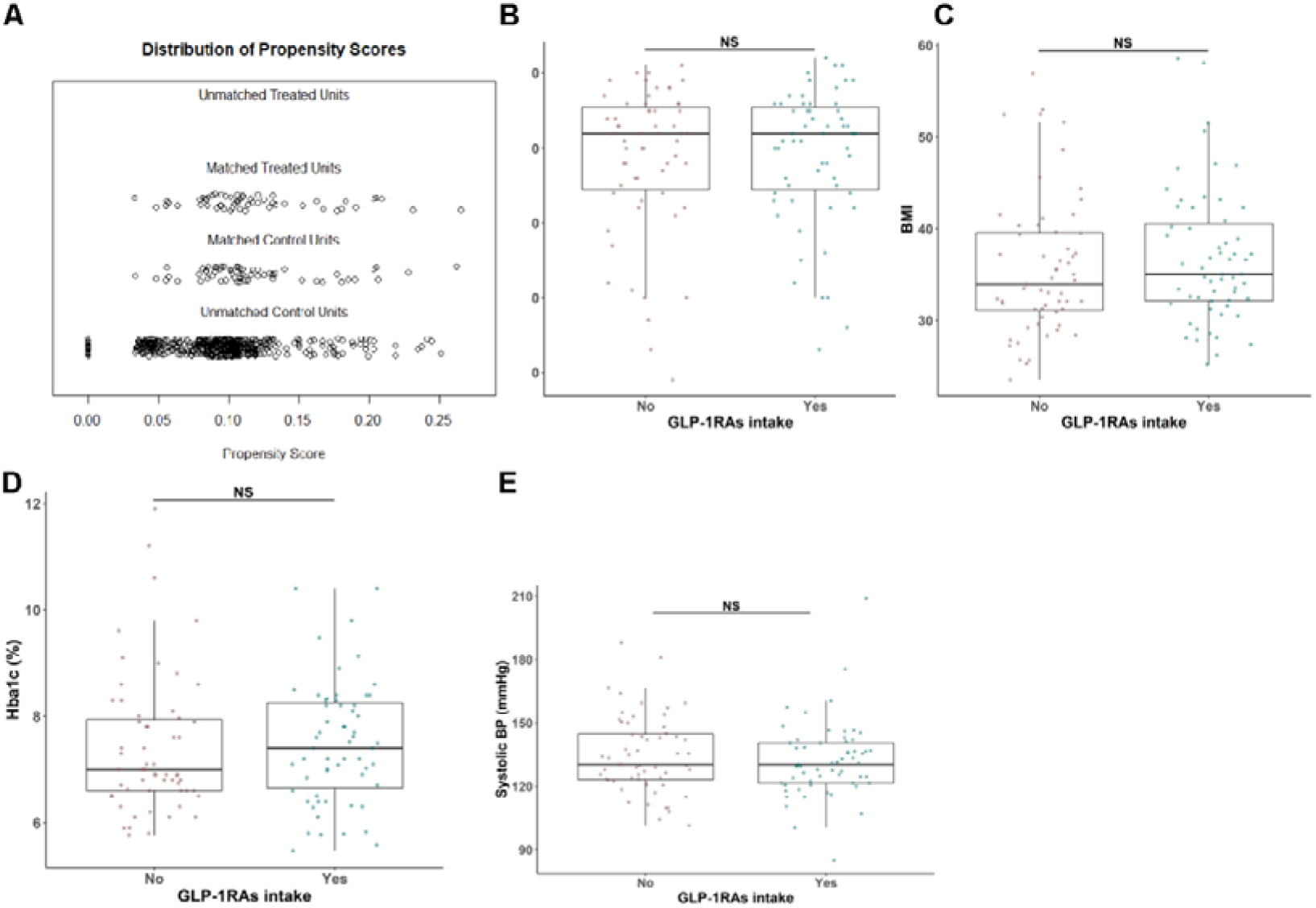
Propensity-score matching of MetaCardis patients with T2D. **(A)** Propensity scores of MetaCardis participants with T2D taking GLP-1 Receptor agonists (GLP-1RAs); N=59) and matched controls (N=59) using nearest neighbors matching with age, sex, disease severity group and hypertension status as covariates **(Suppl.Table.23)**. Comparisons of age **(B)**, BMI **(C)**, glycated hemoglobin **(D)** and systolic blood pressure **(E)**, between MetaCardis patients with T2D diabetics prescribed GLP-1RAs (N=59) and matched controls (as in A; N=59). Mann-Whitney U tests were used to determine statistical significance for B-E. NS: non-significant (*P*>0.05).

## Supplemental Tables

**Supplemental Table 1**. Characteristics of MetaCardis sub-cohorts (BMIS, T2D, IHD).

**Supplemental Table 2**. Spearman correlations between circulating TMAO and biological variables in BMIS.

**Supplemental Table 3**. Spearman correlations between circulating TMAO and clinical variables in BMIS.

**Supplemental Table 4**. BMIS TMAO cluster characteristics.

**Supplemental Table 5**. List of all the variables per Group variable category.

**Supplemental Table 6**. Model hyperparameters and N numbers for each variable group in BMIS.

**Supplemental Table 7**. Explained Variance (EV) of models trained with each variable group in BMIS for each iteration.

**Supplemental Table 8**. Predicted *versus* measured circulating TMAO in BMIS subjects.

**Supplemental Table 9**. Contributions to predictions of the full model by each variable group.

**Supplemental Table 10**. Average SHAP values for all variables for each BMIS subject.

**Supplemental Table 11**. Spearman correlations between circulating TMAO and habitual intake of 37 food categories in BMIS.

**Supplemental Table 12**. Spearman correlations between circulating TMAO and calculated habitual intake of micronutrients in BMIS.

**Supplemental Table 13**. Spearman correlations between circulating TMAO and bacterial species abundance in BMIS.

**Supplemental Table 14**. Differential taxon abundance between high *versus* low TMAO clusters in BMIS.

**Supplemental Table 15**. Model hyperparameters and N numbers for each variable group in T2D.

**Supplemental Table 16**. Model hyperparameters and N numbers for each variable group in IHD.

**Supplemental Table 17**. Explained Variance (EV) of models trained with each variable group in T2D for each iteration.

**Supplemental Table 18**. Explained Variance (EV) of models trained with each variable group in IHD for each iteration.

**Supplemental Table 19**. Average impact on TMAO predictions (SHAP values) for all variables for each T2D subject.

**Supplemental Table 20**. Average impact on TMAO predictions (SHAP values) for all variables for each IHD subject.

**Supplemental Table 21**. Spearman correlations between CAG01909 abundance and intake of drugs in T2D.

**Supplemental Table 22**. Average impact on predicted eGFR (SHAP values) for prescribed medication for MetaCardis T2D subjects.

**Supplemental Table 23**. Characteristics of MetaCardis participants prescribed GLP-1Ras and matched controls.

**Supplemental Table 24**. Materials.

## Methods

### MetaCardis study design and recruitment

Study design, recruitment and exclusion criteria has been extensively described^24,29,34,54^. Patients were subclassified in three groups: BMI-spectrum patients (BMIS^29^; N=837), encompassing MetaCardis participants presenting with metabolic syndrome-related risk factors or conditions (hypertension^55^, obesity^56^ and metabolic syndrome^57^) and patients diagnosed with type-2 diabetes (T2D^58^; N=561) or ischaemic heart disease (IHD; N=356). The IHD group comprised patients with Acute (<15days) Coronary Syndrome (ACS; N=106), Chronic IHD (CIHD; N=157) with normal Left Ventricular Ejection Fraction (LVEF) determined by echocardiography and Heart Failure patients (HF; N=93, LVEF<45%). Ethical approval was obtained from the Ethics Committee CPP Ile-de France, the Ethical Committees of the Capital Region of Denmark (H-3-2013-145) and Ethics Committee at the Medical Faculty at the University of Leipzig (047-13-28012013). Study participants provided written informed consent and the study was undertaken according to Helsinki Declaration-II.

### Sample and phenotypic information collection

Biofluid and biomatter collection has been described elsewhere^24,29,34,54^. Briefly, blood samples were collected in the morning after overnight fasting and fecal samples were collected at home by participants, frozen immediately and transferred to study centers on dry ice within 48h. All samples were stored at -80°C until use. Clinical history, medication and phenotypic information were acquired as described^23,28,34,54^ with standardized procedures across centers. Participants reported habitual food intake through a customized Food Frequency Questionnaire (FQQ)^59^. Bioclinical variables were measured in a single center according to standard procedures^28^. Estimated glomerular filtration rate (eGFR) was calculated with the CKD-EPI formula without ethnicity adjustment^61^.

### Metabolic profiling

#### ^1^H-Nuclear Magnetic Resonance (^1^H NMR) spectroscopy

Spectra acquisition, using an Avance spectrometer (Bruker) at 600 MHz; and structural assignments have been extensively described previously^25,29^. Absolute quantifications were derived using the “*In Vitro* Diagnostics for research” (IVDr) quantification BI-LISA and BI-Quant algorithms (Bruker).

#### Gas Chromatography coupled Mass Spectrometry (GC-MS)

Serum samples (100μl) were prepared, analyzed and processed as described^29,35^. Briefly, protein was methanol-precipitated, methanol was evaporated to dryness and subsequent to derivatization samples were injected to an Agilent 7890B-5977B Inert Plus GC-MS system. The chromatographic column was an Agilent ZORBAX DB5-MS (30m x 250μm x 0.25μm + 10m Duragard). The temperature gradient was 37.5min long and the mass analyzer was operated in full scan mode between 50 and600 m/z. Peaks were annotated with the use of the Fiehn library (Agilent G1676AA Fiehn GC/MS Metabolomics RTL Library, User Guide, Agilent Technologies, https://www.agilent.com/cs/library/usermanuals/Public/G1676-90001_Fiehn.pdf). Metabolic features with low reproducibility or linearity were removed from the dataset, resulting in 102 annotated metabolic features.

#### Ultra-Performance Liquid Chromatography-Tandem Mass Spectrometry (UPLC-MS/MS)

UPLC-MS/MS performed on a Waters Acquity UPLC-Xevo TQ-S UPLC-MS/MS system equipped with an Acquity BEH HILIC (2.1×100mm, 1.7μm) chromatographic column was employed to determine TMA, TMAO, choline, betaine, γ-butyrobetaine, betaine aldehyde, butyryl-carnitine, isovaleryl-carnitine, OH-isovaleryl-carnitine, stearoyl-carnitine, oleoyl-carnitine, linoleoyl-carnitine, myristoyl-carnitine, lauroyl-carnitine and decanoyl-carnitine as described previously^28,34^.

TMAO circulating values of all MetaCardis participants were log_10_-transformed and subsequently the median was subtracted and divided by the standard deviation^21^ (SD; regularized TMAO values thereafter).

#### Metagenomic analysis

Phylogenetic microbiota profiles were built after correction for bacterial load as extensively described^24,29,35,54^ using a protocol devised by Vandeputte *et al*^61^ with modifications. Briefly, total faecal DNA was extracted following the International Human Microbiome Standards (IHMS) guidelines (SOP 07 V2 H) and samples were sequenced using ion-proton technology (ThermoFisher Scientific). Gene abundance profiling was performed using the 9.9 million gene integrated reference catalog of the human microbiome, as described^24,29,34,54^.

#### Customized microbial module analysis (GMM)

Manually-curated customized module sets focusing on anaerobic bacterial and archaeal fermentation processes relevant to the human gut microbiota were assembled as previously extensively described^24,29,34,54^.

#### Statistical analyses

Statistical analysis was undertaken using R (v4.03)^62^. For comparing two groups we used Mann-Whitney U and for multiple group comparisons Kruskal-Wallis tests. Unadjusted Spearman correlations were computed using R, whilst adjusted Spearman correlations with ppcor (v1.1). *P*-values were corrected for multiple comparisons using the Benjamini-Hochberg method.

### Machine Learning (ML) analysis

#### Variable groups

Patient phenotypic variables were separated into 10 groups **(Suppl.Table.5)**. Specifically, biological parameters included biochemical and clinical serum laboratory tests including lipids, glycated haemoglobin, creatinine and eGFR. Clinical parameters consisted of clinical history, BMI, systolic and diastolic blood pressure, anthropometric variables and stool frequency and type. Demographic information comprised age, physical activity, educational and income levels, smoking status, ethnicity and country of recruitment. Drug variables included intake of common medication as described^34^, number of antibiotic courses in the last 5years and number of antihypertensive, antidiabetic and lipid-lowering treatments. Dietary parameters included habitual consumption of 37 food items, daily nutrients intake derived from these food items calculated as in^59^, alternative Healthy Eating^63^ (aHEI), Dietary Approaches to Stop Hypertension^64^ (DASH) and Dietary Diversity^65^ (DDS) scores. Clinical risk factors variables included age, systolic and diastolic blood pressure, glycated haemoglobin levels, fasting cholesterol levels, smoking status and waist circumference as described^29^. Serum metabolomics comprised the absolute or relative levels of 116 circulating metabolites determined by GC-MS or UPLC-MS/MS^29,34^. Urine metabolomics included absolute quantification of 47 urine metabolites from ^1^H-NMR spectra calculated with the IVDr algorithm. Microbiota variables included abundance of 699 bacterial species present in at least 20% of MetaCardis patients corrected for microbial load and the first 10 principal components from a PCA of relative microbial gene abundances^29,34^. Microbiome (Modules) group included abundance of 116 manually curated bacterial modules^29^. In all cases categorical variables were converted into dummy variables using caret (v6.0.86).

#### Boosted decision trees (Xgboost)

We predicted regularized circulating TMAO levels by using gradient boosting decision trees based on the xgboost algorithm (v1.3.2.1)^66^, co-opting a strategy from Bar *et. al*^21^. Xgboost consistently outperforms other algorithms in Kaggle competitions for tabular data. For each of our 10 variable groups **(Suppl.Table.5)** we optimized xgboost models using 5-fold cross-validation and two sequential hyperparameter grids searches (972 different parameter combinations for each feature group in total) to predict mean-centred and unit variance-scaled (regularized) TMAO levels in the left-out group using root-mean-square error (RMSE) to evaluate model outcomes. After parameter optimization, we predicted circulating TMAO using 5-fold cross-validation for 100 iterations using as input the variables of each group or all variables (full-model). For each round, we calculated the coefficient of determination using the rsq function from yardstick (v0.0.7) and the predicted regularized TMAO values. Xgboost models trained with 80% of each patient group participants during cross-validation (5 for each round) were saved and used for feature attribution analysis in the left-out group (see below).

#### SHapley Additive exPlanations (SHAP) analysis

We interpreted our ML models and assigned relative importance to variables influencing circulating TMAO levels by co-opting SHAP values, as expanded for tree-based ML models^26^ and recently used to objectively evaluate factors driving metabolite plasma levels in humans^21^. Briefly, for each prediction the SHAP value of a particular variable is the difference in the model output when this variable is included *versus* when excluded. Variables are added to the model in all possible orderings and the SHAP value is computed from the average of model outcomes^26^. Variable SHAP values for each individual in the 5 left-out groups were extracted from corresponding xgboost models trained on the remaining 80% participants for each variable group using SHAPforxgboost (0.1.0) and averaged over 100 iterations. We assigned relative importance to each variable by computing the mean of the absolute SHAP value for all individuals in each disease group similarly to Bat *et al*.^21^. For swarm-plots depicting each individual participants SHAP values, variable values were inverse-normalized using the RNOmni (v1.0.0) package.

#### Clustering

Clustering was performed using the built-in R k-means function with the Hartigan-Wong algorithm using 25 random sets, as described^25^.

#### Multivariate analysis

To identify microbiota composition differences between individuals split in TMAO clusters we performed multivariate homogeneity analysis of Bray-Curtis dissimilarity matrices at the species level (699 species present in at least 20% of our population) and determined statistical significance with a permutation ANOVA test (999 iterations) using the vegan (v2.5.7) package. Permutation analysis of variance (PERMANOVA) of regularized circulating TMAO versus Bray-Curtis taxonomic dissimilarity matrices with age, sex and country of recruitment as co-variates for 999 iterations were performed with vegan (v2.5.7).

#### Mediation analysis

We performed mediation analysis^67^ to assess the putative impact of eGFR on plasma TMAO increase with age and conversely the impact of TMAO on eGFR decline with age. We used the Preacher and Hayes bootstrapping method, as implemented in the mediation (v4.5.0) R package^68^, using general linear models with sex and country of recruitment as covariates. Confidence intervals and Bayesian *P* values were computed after 999 simulations.

#### Propensity score matching

MetaCardis participants with T2D were propensity-score matched using the R package MatchIt^69^ (v.4.1.0) with age, sex, disease group and hypertension status as covariates using nearest neighbor matching determined by generalized linear models. All covariates were given equal weight.

#### Preclinical models

A complete list of antibodies and reagents used in these studies are appended (Suppl.Table.24)

#### *In vitro* experiments

##### Cell Culture and Protein Extraction

Primary human adult kidney fibroblasts from a single donor (Cambridge Bioscience) were cultured in 10% FCS low-Glucose DMEM with 1% Pen/Strep antibiotics (Sigma). For acute (up-to 30min) TMAO stimulation, cells (100,000/condition) were serum-starved for 1h in a physiologic serum-free buffer as previously described^43^. Fibroblasts were preincubated with Tramenitib (10nM, Shelleckchem), BAPTA-AM (20μM, Molecular Probes) or vehicle for 30min before TMAO (Sigma) stimulation. For longer-term (24h) stimulation, fibroblasts were in serum-free low-glucose DMEM overnight and subsequently stimulated with TMAO, TGF-β1 (5ng/ml, R&D), or their combination. At the end of the experiment fibroblasts were lysed and stored at −80°C until further use, as described^42^.

##### Cytosolic [Ca^2+^] Measurements

Fibroblasts were serum-starved overnight in low-glucose DMEM and subsequently loaded with 5μM fura-2-AM (Molecular Probes) in pH 7.4 Hanks Balanced Salt Solution (HBSS; Sigma) containing Ca^2+^ and Mg^2+^. Measurements were obtained on an epifluorescence inverted microscope equipped with a✉×20 fluorite objective. Single cell intracellular Ca^2+^ ([Ca^2+^]_i_) was monitored using excitation at 340 and 380nm, through a monochromator (Cairn Research). Emitted light was reflected through a 515nm filter to a QImaging Retiga CCD camera (Cairn Research) and digitized to 12-bit resolution. All imaging data were collected and analyzed using software from Andor.

##### Animal Procedures

All animal experiments were conducted in accordance with the United Kingdom Home Office Animals 1986 Scientific Procedures, with local ethical committee approval (Project License 70/8356). Male C57BL/6J mice (Charles River) at 6-8weeks of age were fed a control, 0.12% w/w TMAO- or 1% w/w choline-containing diets (Teklad; 6animals/group) for 6weeks^18^. Subsequently, Unilateral Ureter Obstruction (UUO) was established as previously described^42^. Briefly, mice were anesthetized with isoflurane and the abdominal cavity was exposed using midline laparotomy, the right ureter was isolated and tied off 0.5cm from the pelvis. The left ureter was left unclamped and served as the sham-operated control. At day 5 the kidneys were harvested. Both kidneys from each animal (UUO and sham) were cut in half longitudinally. One half of each kidney was snap-frozen in liquid nitrogen and subsequently stored at −80°C for Western blotting. The other half was fixed with 10% Formalin (Sigma) for 16h at 4°C, transferred to a 70% v/v ethanol solution for a further 24h and was then paraffin-embedded for immunohistochemistry.

##### Western Blot

We used the NuPAGE electrophoresis and buffer system (Invitrogen) for immunoblot analysis of kidney samples or cellular lysates as described^40,41^. Proteins were visualized with ECL Prime (GE Healthcare). Optical densities of bands of interest were determined using ImageJ 1.46r (NIH) and normalized against loading controls. The value of the normalized control sample was arbitrarily set to 1. Membranes were stripped using the Restore Plus reagent (Fisher Scientific), and re-probed with appropriate loading controls.

##### Immunohistochemical Staining

Formalin-fixed kidneys were embedded in paraffin and 4μm sections were cut by the Barts Cancer Institute Pathology Unit. Staining for Sirius red, α-Smooth Muscle Actin (αSMA) and F4/F80 were performed as described^42^ with the DISCOVERY XT (Ventana) automated slide processing instrument using the OmniMap reagents (Ventana), according to the manufacturer’s recommendations. Images were captured at ×20 magnification, using a Zeiss AxioPhot microscope with an AxioCam HRc camera. Five kidney cortex images were captured per mouse and staining was quantified as percentage of total area, using ImageJ 1.46r (NIH).

